# BanffNET, a Deep Learning System for Comprehensive Histological Lesion Quantification in Kidney Transplant Biopsies

**DOI:** 10.64898/2026.08.28.26360029

**Authors:** Giorgio Buzzanca, Chiara Pala, Junling He, Rianne Hofstraat-Boersma, Alessandra Tammaro, Dominique van Midden, Roman Bülow, David L. Hölscher, Anja S. Mühlfeld, Maximilian Koeller, Nicolas Kozakowski, Georg A. Böhmig, Philip F. Halloran, Danny van der Helm, Soufian Meziyerh, Jan-Hendrik Venhuizen, Saskia Haitjema, Jouke Dijkstra, Luuk B. Hilbrands, Eric J. Steenbergen, Arjan D. van Zuilen, Azam S. Nurmohamed, Frederike J. Bemelman, Imke B. Bruns, Giulia Callegaro, Bob van de Water, Tobias T. Pieters, Gerben E. Breimer, Giovanni M. Rossi, Enrico Fiaccadori, Umberto Maggiore, Joris J.T.H. Roelofs, Francesca Testa, Francesco Fontana, Adeyemi Adefidipe Abiola, Marco Delsante, Garry L. Corthals, Hessel Peters-Sengers, Tri Q. Nguyen, LUMC Global Kidney Biopsy Reader Group, Priyanka Koshy, Sandrine Florquin, Peter Boor, Y.K. Onno Teng, Maarten Naesens, Aiko P.J. de Vries, Jesper Kers

**Author notes:** Corresponding author: Jesper Kers, MD, PhD.

## Abstract

Accurate, reproducible interpretation of kidney allograft biopsies is critical for the diagnosis of graft injury and for informing prognosis and clinical management. The international Banff classification is a consensus diagnostic system based on semiquantitative histological lesion scoring according to either lesion extent or severity in kidney transplant biopsies. However, pathologist scoring is limited by interobserver variability, constrained scalability, and the inherent nature of the scoring system itself. Here we present BanffNET, a weakly supervised, probabilistic deep learning framework that combines self-supervised feature extraction with a novel Bayesian multiple-instance learning framework to predict (continuously) the full spectrum of Banff lesion scores directly from whole-slide images (WSIs). Using lesion-specific aggregation functions tailored to localized (modeling severity) and diffuse histological lesions (modeling extent), BanffNET generates interpretable, patch-level probability maps and calibrated slide-level scores. BanffNET’s performance was assessed relative to consensus, biological correlates of rejection and clinical outcome, demonstrating superior consistency, transportability and generalization. Trained on 7,533 WSIs from three cohorts, BanffNET demonstrates consistent performance on 12,687 WSIs across five external validation cohorts, matching or surpassing individual expert pathologists across lesion assessments. BanffNET scores align more closely than pathologist Banff scores with molecular profiles of rejection, offering an objective, transparent, biologically grounded framework for computational pathology with relevance beyond kidney transplantation.

## Introduction

Histological evaluation of tissue biopsies is essential to medicine, providing critical insights into disease processes, prognosis, and informing therapeutic decisions. In kidney transplantation, such evaluations are guided by the Banff classification^1^, a globally adopted framework that synthesizes histological lesion scores with serological and immunohistochemical findings to diagnose and classify graft rejection. This classification is based on semiquantitative lesion scoring conducted by pathologists across various renal compartments^2^.

Despite its clinical value, applying the Banff classification demands extensive expertise and can be limited by significant interobserver variability. The complex, semi-quantitative nature of pathologists’ Banff lesion scores and the patchy, often subtle presentation of key features pose challenges for reproducibility and scalability, leading to diagnostic uncertainty and potential variability in patient management. Previous studies underscored the inherent subjectivity in pathologists’ Banff lesion score evaluation and the need for standardization, training, and diagnostic companion tools^3,4^.

As diagnostic workflows increasingly incorporate digital pathology, there is a growing interest in leveraging artificial intelligence (AI) to support and standardize histological assessments. A significant advancement in this domain is the development of histology foundation models (HFM), deep learning systems trained on large-scale, unannotated whole-slide images using self-supervised learning (SSL) techniques. These models have demonstrated the ability to learn rich, generalizable visual representations from histological slides, serving as a robust foundation for downstream diagnostic tasks^5,6^. Domain-specific SSL approaches have proven particularly effective in histopathology, in which incorporating pathological priors during pre-training enhances model performance across tissue types and clinical applications^7^. However, many existing AI applications simplify complex pathology into discrete classification outputs, limiting interpretability, and often underperforming if pathology is subtle, multifactorial, or spatially heterogeneous as is often the case in kidney transplant pathology.

In this study, we introduce a novel suite of deep learning algorithms called **BanffNET**. BanffNET leverages a combination of HFM feature extraction^8^ with probabilistic graphical modeling to support fully quantitative and explainable lesion analysis in kidney transplant biopsies. The method was designed to work under weak supervision, requiring only slide-level labels, and incorporates a low-capacity Bayesian multiple-instance learning (MIL) structure that enhances generalization and interpretability^9^. By generating probabilistic scores for each lesion at the patch level and aggregating them to slide-level outputs (BanffNET scores), the system aligns closely with the principles of expert histological review by design^10^, while allowing granular quantifications of kidney transplant lesions at scale.

## Results

### Training and validation cohorts and sample characteristics

BanffNET was trained on a heterogeneous multicenter cohort of a total of 7,533 whole slide images (WSIs) of post-transplant biopsies from three centers in The Netherlands (Amsterdam, Leiden and Utrecht) and externally validated on 12,687 WSIs from five kidney transplant centers across Europe (Leuven/Belgium, Vienna/Austria, Aachen/Germany, Nijmegen/The Netherlands and Parma/Italy), representing substantial clinical and technical variation in terms of case mix, clinical biopsy protocols, scanners, staining procedures, and hospital workflows. Beyond the 5 external validation cohorts, an international multi-reader study was conducted that included 67 international pathologists, who independently scored 36 biopsies^11^. This allowed us to compare BanffNET lesion scores in the context of inter-observer variation. A detailed description of the multi-reader study cohort can be found in the **Online Methods,** and an overview of baseline characteristics can be found in **Extended Data Table 1**. Biopsies were assessed for 15 Banff- defined lesions using three standard staining protocols: hematoxylin and eosin (HCE), Jones’ silver (silver), and periodic-acid Schiff (PAS). The 15 lesions were annotated according to the most recent Banff classification criteria, as described in Roufosse et al.^2^ and Naesens et al.^1^, and include:

- *Active inffammatory lesions*: glomerulitis (g), peritubular capillaritis (ptc), intimal arteritis/vasculitis (v), total cortical inflammation (ti), interstitial inflammation in non-scarred areas (i), interstitial inflammation in scarred areas (i-ifta), tubulitis in non-scarred areas (t) and tubulitis in scarred areas (t-ifta);
- *Chronic fibrosing lesions*: glomerular basement membrane double contours (cg), mesangial matrix increase (mm), interstitial fibrosis (ci), tubular atrophy (ct) (grouped together as [ifta]), arteriolar hyalinosis (ah) and vascular intimal thickening (cv);
- *Damage-associated lesions*: acute tubular injury (ati) and thrombotic microangiopathy (tma).

Additional models were trained to predict the percentage of glomeruli affected by focal segmental glomerulosclerosis (fsgs) and global glomerulosclerosis (gs), important chronic lesions, despite not being officially included in the Banff classification. An overview of the pathologist Banff lesion score distributions can be found in **Extended Data Table 2**. A visual overview of the image data can be found as random patch mosaics in **Extended Data Figures 1 and 2** and **Extended Data Figure 3** visualizes the variability in color distributions.

### Probabilistic multiple instance learning with Bayesian network modeling in BanffNET

BanffNET integrates Bayesian principles and introduces a novel MIL aggregation scheme based on the Noisy-OR logical operator, representing the first use of such probabilistic modeling in conjunction with self-supervised learning in computational pathology. In short, gigapixel WSIs were tessellated into image patches measuring 256×256 pixels. UNI^12^, a general-purpose histology foundation model pre-trained with DINOv2^13^ on >100 million histology images across 20 tissue types, was used to convert image patches into patch-level feature vector representations. Slide-level semi-quantitative (ordinal) pathologist Banff lesion scores were converted into probabilities, thereby framing the problem as a probabilistic regression task (**Extended Data Table 3)**. Pathologist Banff lesion scores represent either the *extent* of a lesion, i.e. what percentage of the kidney is affected (e.g. interstitial inflammation) or the *severity* of a lesion, i.e. the maximum number of inflammatory cells found within a structure (e.g. tubulitis). Focal lesions representing maximum severity were modeled using Noisy-OR Instance Pooling (NIP), while diffuse lesions representing spatial extent were modeled using Gated Instance Pooling (GIP). Mathematical definitions and additional details are provided in the **Online Methods**. The final BanffNET system comprises 51 individual GIP or NIP models, corresponding to 17 lesions assessed in each of three routine stains: HCE, silver, and PAS. For four selected lesions (ati, ifta, ptc and t), the GIP/NIP performance was compared with state-of-the-art (larger) multiple instance learning frameworks, ABMIL and TransMIL, indicating increased performance for the GIP/NIP models, despite using over 2,500-fold fewer model parameters (**Extended Data Table 4; Extended Data Table 5**), leading to a significantly more ecologically sustainable solution. On a single NVIDIA Quadro RTX 6000 (24GB VRAM), it takes on average 18 seconds to process the 17 BanffNET models per WSI end-to-end (i.e. 5 hours for 1,000 WSIs). Although the absolute differences in performance were modest, BanffNET offered more than just a numerical advantage over the attention-based MIL architectures. For each of the GIP/NIP models, high-resolution *pixel-level* explainability maps were then generated using the Generic Attention Explainability (GAE) method^14^, applied to the full architecture consisting of the frozen UNI ViT backbone and the lesion-specific BanffNET heads (see **Online Methods**). Its probabilistic modeling framework produced interpretable probability maps that corresponded closely with established lesion patterns, offering localized visual explanations that were more precise and anatomically coherent than those generated by current state-of-the-art attention-based methods. In a representative case with peritubular capillaritis (ptc), BanffNET’s *pixel-level* explainability map highlighted lesion regions in line with expert assessment, whereas the *patch-level* attention maps from attention-based multiple instance learning models were more diffuse and diagnostically ambiguous (**Extended Data Figure 4**).

An overview of the BanffNET computational workflow, examples of pixel-level visualizations, and a radar plot summarizing the results from clinical, molecular and histological validation can be found in **Figure 1**.

**Figure 1.**
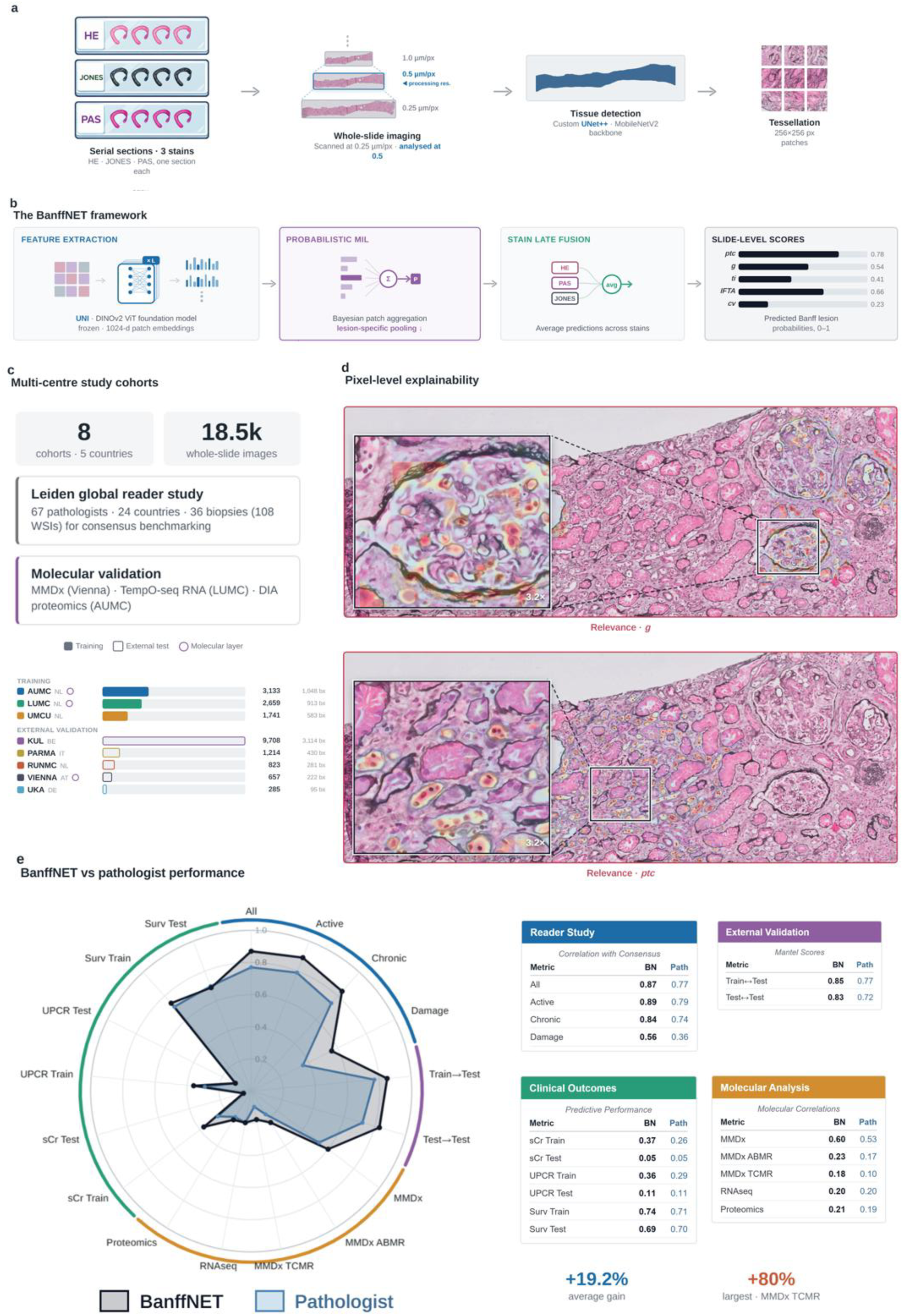
The BanffNET system and its validation strategy. **a.** For each kidney transplant biopsy, three serial sections were stained (HCE, Silver and PAS), digitized by whole-slide imaging at 0.25 µm/pixel and analyzed at 0.5 µm/pixel. Diagnostic tissue is delineated by a custom in-house UNet++ segmentation model with a MobileNetV2 backbone, and the masked region is tessellated into non-overlapping 256×256-pixel patches. **b.** Each patch is encoded into a 1,024-dimensional feature vector by the frozen UNI^8^ histology foundation model (a ViT^15^ pretrained with self-supervised DINOv2^13^ on >100 million histology images). Lesion-specific probabilistic multiple-instance learning heads aggregate patch-level probabilities into slide-level lesion scores - Noisy-OR Instance Pooling (NIP) for focal lesions and Gated Instance Pooling (GIP) for diffuse, proportion-based lesions. Stain-specific predictions are combined via late fusion to yield the final biopsy-level BanffNET scores. **c.** Overview of the eight multi-center cohorts, indicating the number of slides contributed by each center and the orthogonal data modalities available for biological validation. **d.** Representative pixel-level relevance maps generated by Generic Attention Explainability (GAE), providing anatomically coherent, lesion-specific localization of model predictions. Upper panel: glomerulitis (BanffNET g score); lower panel: peritubular capillaritis (BanffNET ptc score) **e.** Radar plot summarizing BanffNET performance results across all 15 histological, molecular, clinical, and cohort transportability validation tasks (axes scaled from 0 to 1). The average gain for BanffNET compared with pathologist assessment was 19.2% with a maximum gain of 80% for MMDx TCMR archetype retrieval.

### BanffNET predictions correlate with pathologist-assigned lesion scores across datasets

BanffNET generates continuous lesion scores in kidney transplant biopsies, enabling quantitative assessment across the full spectrum of lesion severity. In the training cohort, BanffNET continuous lesion scores showed strong and consistent correlation with semiquantitative histological Banff lesion scores (all P<0.0001), with clear visual incremental separation across the levels in all lesions assessed (**Extended Data Figure 5**; **Extended Data Table 6**). These correlations between BanffNET lesion scores (calculated with frozen model weights) and pathologist Banff scores were preserved in the validation cohorts (all P<0.001 for trend across binary, ordinal or continuous scores), (**Extended Data Figure 6; Extended Data Table 6**). These results highlight the model’s generalizability across diverse data sources and suggest that its scores preserve clinically meaningful stratification across histological score categories without relying on threshold-based interpretation, despite significant domain shift due to staining variation and pathologist interobserver variability.

Post-hoc discrepancy analysis identified several cases labeled as v3 arteritis by the pathologist that received low BanffNET v scores. Detailed multi-reader central review of these WSIs (n = 8) identified alternative pathological processes such as cortical necrosis, acute tubular necrosis, or thrombotic microangiopathy as the dominant findings, and 7 of 8 cases could retrospectively be reclassified to v0, thus representing initial errors in the histological labeling rather than aberrant performance of the BanffNET models (**Extended Data Table 7**). To further investigate discrepancies between continuous BanffNET and semiquantitative pathologist Banff lesion scores more systematically, the 10 validation cases with the largest divergence between BanffNET and pathologist Banff lesion scores, defined as maximum Euclidean distance, were blindly reviewed by a panel of three nephropathologists (**Extended Data Tables 8–17; Extended Data Figure 8**). In half of the cases, discordance was attributable to technical artifacts, such as poor stain quality, focus issues, or WSIs with mislabeled stain types, potentially impacting model predictions. In the remaining cases, disagreement stemmed from complex or overlapping pathology, including infection-related injury (pyelonephritis), extensive glomerulosclerosis, or recurrent glomerulonephritis.

### BanffNET aligns with pathologist consensus in a global multi-reader study

We next evaluated BanffNET’s performance against expert *consensus* review. We conducted an international reader study involving 67 renal pathologists who independently scored 36 kidney transplant biopsies^11^ (see **Online Methods**). To mitigate the interobserver variability in the pathologist Banff lesion scoring, we used the mean Banff lesion scores across 67 experts as a proxy consensus to compare with the BanffNET predictions. This mean consensus score enabled a more granular evaluation of lesion severity than semiquantitative scores and captured subtle diagnostic features that may be recognized only by more experienced readers. BanffNET scores showed high correlations with mean consensus scores across lesions (**Figure 2a**). We next assessed to what extent BanffNET and individual pathologists aligned with the mean consensus score for each of the Banff lesions by Pearson correlation. BanffNET lesion scores consistently aligned with the mean consensus scores to a degree comparable to the top-performing individual pathologists (**Figure 2b**), independent of years of experience. For certain lesions, most notably total inflammation (ti) and peritubular capillaritis (ptc), BanffNET outperformed all individual experts in terms of correlation with the mean consensus scores. These findings illustrate BanffNET’s potential to match or exceed expert-level performance, including in lesions known to be prone to interpretative variability. For example, we identified a case where BanffNET predicted a high score for intimal arteritis (v) whereas 77% of pathologists scored this lesion as absent (v0). However, CD45-CD34 double immunohistochemical staining (**Figure 2c**), respectively identifying leukocytes and endothelium, clearly demonstrated leukocyte infiltration beneath the endothelium, consistent with intimal arteritis. In a second case, BanffNET predicted a high score for glomerular double contours (cg) whereas 95% of pathologists scored this lesion as absent (cg0). Electron microscopy (EM), however, identified early double contours (**Figure 2d**), which according to the Banff classification are classified as *cg1a*: not visible on light microscopy but only identified with EM^2^. This particular patient was known to have antibody-mediated rejection diagnosed 3 months earlier, not responding to anti-rejection treatment.

**Figure 2.**
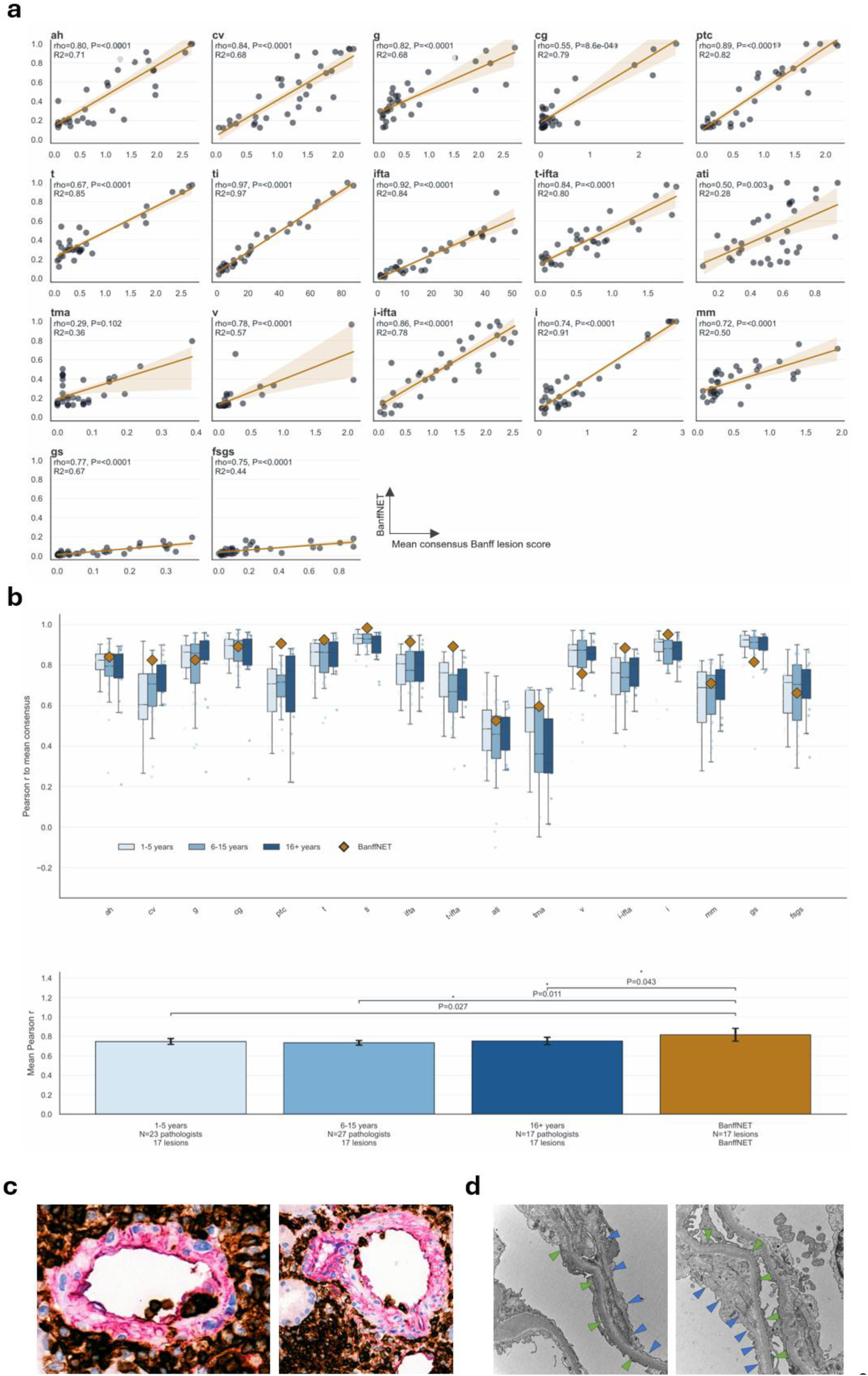
Correlation of BanffNET scores to the consensus of 67 pathologists from the Leiden Global Reader Study. **a.** Correlation of BanffNET lesion scores with the mean consensus scores by the 67 renal pathologists. **b.** Comparison of the BanffNET lesion scores and the individual pathologist histological Banff lesion scores against the consensus of all 67 readers. Across all lesions, BanffNET showed stronger correlations with the consensus than individual pathologists, independent of years of experience (all p<0.05, one-sided Welch t-test testing superiority of BanffNET vs individual pathologists). **c.** Case from the reader study where the majority of readers (77%) scored absence of intimal arteritis (v0), whereas BanffNET scored high for intimal arteritis. Double staining for CD45 (leukocytes, brown) and CD34 (endothelium, red) demonstrates leukocyte infiltration beneath the endothelium, consistent with intimal arteritis. **d.** Case from the reader study where most readers (95%) scored absence of double contours (cg0), whereas BanffNET scored high for glomerular double contours (cg). Electron microscopy shows the original glomerular basement membrane (green arrowheads) and the newly formed glomerular basement membrane (blue arrowheads), indicating early double contours only visible by EM (cg1a).

### BanffNET captures clinically meaningful correlations between histological lesions associated with rejection phenotypes

The pathologist Banff scores are not independent; rather, they reflect biologically interrelated processes, particularly in the context of rejection phenotypes (**Figure 3a-b**). Although BanffNET was trained to predict each lesion independently, without explicit modeling of inter-lesion dependencies (a multitasking objective was purposely not deployed during training), we assessed whether the system implicitly recapitulates the population-based correlation structure seen in pathologist annotations. BanffNET reliably reproduced the associations between lesions (**Figure 3c-d**). On the validation cohorts strong correlations were observed among lesions associated with microvascular injury and chronic damage, including peritubular capillaritis (ptc) and glomerulitis (g), ptc and chronic glomerulopathy (cg), as well as glomerulitis and thrombotic microangiopathy (tma).

**Figure 3.**
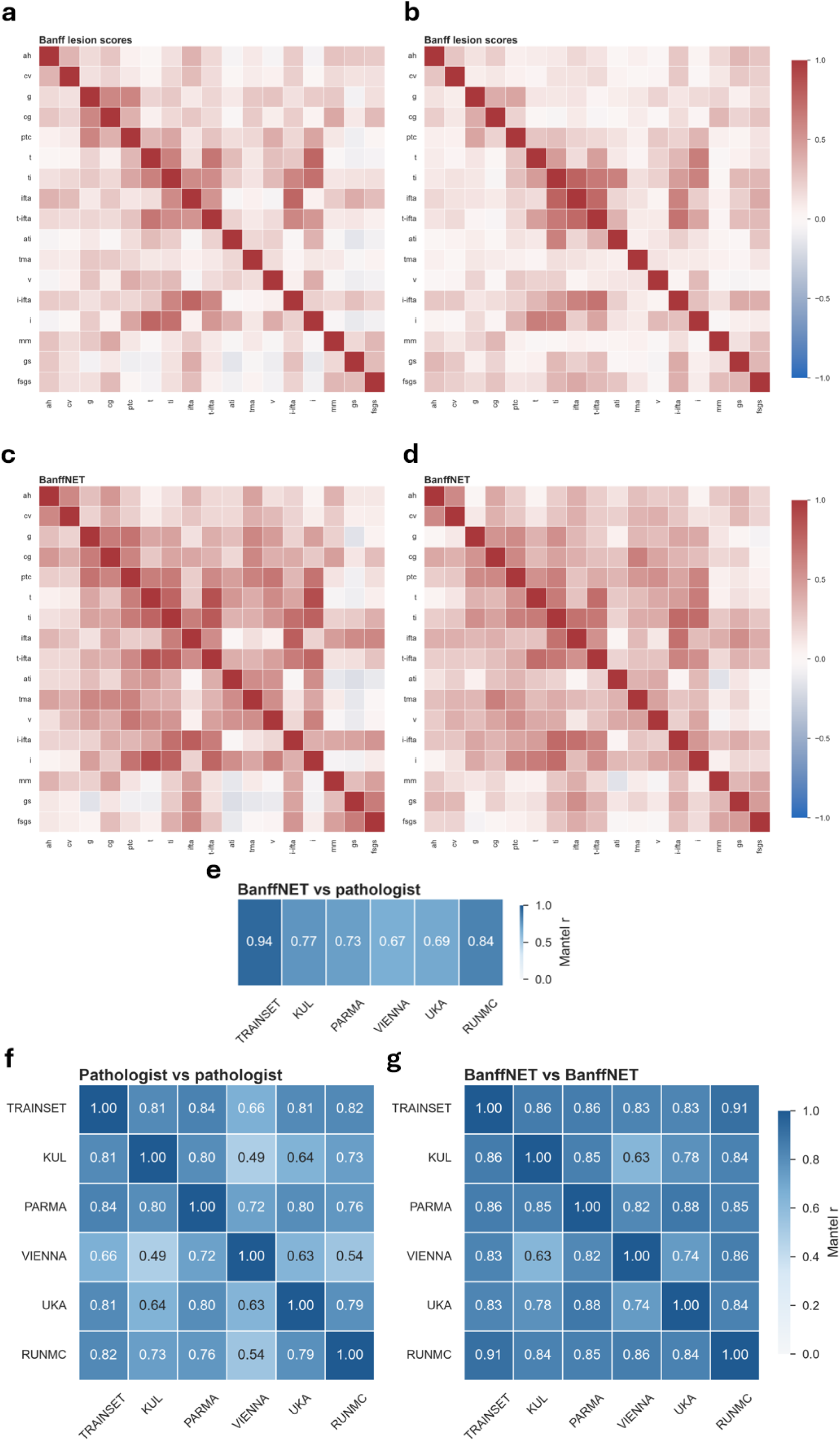
Intra- and inter-cohort lesion correlations for Pathologist Banff scores and automated BanffNET scores. **a.** Training set lesion correlation matrix of pathologist Banff scores. **b.** Validation set lesion correlation matrix of pathologist Banff scores. **c.** Training set lesion correlation matrix of BanffNET scores. **d.** Validation set lesion correlation matrix of BanffNET scores. **e.** Mantel statistics for within-cohort consistency between pathologist Banff scores and BanffNET scores. **f.** Pairwise Mantel statistics for between-cohort consistency of pathologist Banff scores. **g.** Pairwise Mantel statistics for between-cohort consistency of BanffNET scores.

To compare how BanffNET and pathologists captured the interrelationships between lesions, we used the Mantel test, a method for comparing similarity patterns between matrices. A higher Mantel statistic indicates greater agreement between predicted lesion interdependencies and those seen in expert scoring. Within-cohort comparison (**Figure 3e**) showed high matrix consistency in the training cohort (Mantel statistic = 0.94), which was preserved in the validation cohorts (Mantel statistic range 0.67 – 0.84). Across cohorts, the average Mantel statistic for pathologist Banff score-derived correlation matrices was 0.71 (range 0.49 - 0.84), compared to 0.83 (range 0.63 - 0.91) for matrices based on individual pathologist Banff lesion scores, paired-sample t-test P=0.0001 (**Figure 3f–g**). This indicates that BanffNET not only produces accurate individual lesion scores, but also better preserves the broader structure of histopathological relationships across kidney transplant biopsies compared with individual pathologist assessments.

### BanffNET lesion scores associate with kidney function markers and graft outcome

To further assess clinical relevance, we modelled BanffNET lesion scores against two indicators of graft integrity, urinary protein-to-creatinine ratio (UPCR) and serum creatinine (SCr), using cross-sectional analyses. Multivariable linear regression models incorporating BanffNET lesion scores consistently showed higher (training cohort) or equal explained variation (validation cohort) for UPCR and SCr compared to those using pathologist Banff lesion scores (**Extended Data Figure 8**). Univariable associations of BanffNET and pathologist Banff scores with both outcomes are reported in **Extended Data Figure 9**.

Cox models for death-censored graft failure, trained using the Amsterdam, Leiden, and Utrecht cohorts and evaluated in the Leuven and Vienna cohorts, showed that BanffNET-based multivariable models offered equal or higher overall discrimination for death-censored graft failure than pathologist Banff score-based multivariable models, with higher c-indices on the training cohort and equal c-indices on the validation cohorts (**Figure 4a**). Time-dependent ROC analysis further indicated similar cumulative-dynamic AUC values for multivariable BanffNET and pathologist Banff score models throughout follow-up on the training and validation cohorts (**Figure 4b**), with only minor differences mostly in the train cohort and for those graft failures closest to the time of biopsy (**Extended Data Figure 10**). Patient stratification into low-, intermediate-, and high-risk groups based on tertiles of the multivariable death-censored graft failure risk score showed comparable stratification between BanffNET- and pathologist Banff score-based models in both the training and validation cohorts (**Figure 4c**). Across all models, a higher proportion of patients fell into the low- and intermediate-risk strata, reflecting the large number of protocol biopsies in the validation cohort. Univariable associations of BanffNET and pathologist Banff scores with death-censored graft failure are reported in **Extended Data Figure 11**.

**Figure 4.**
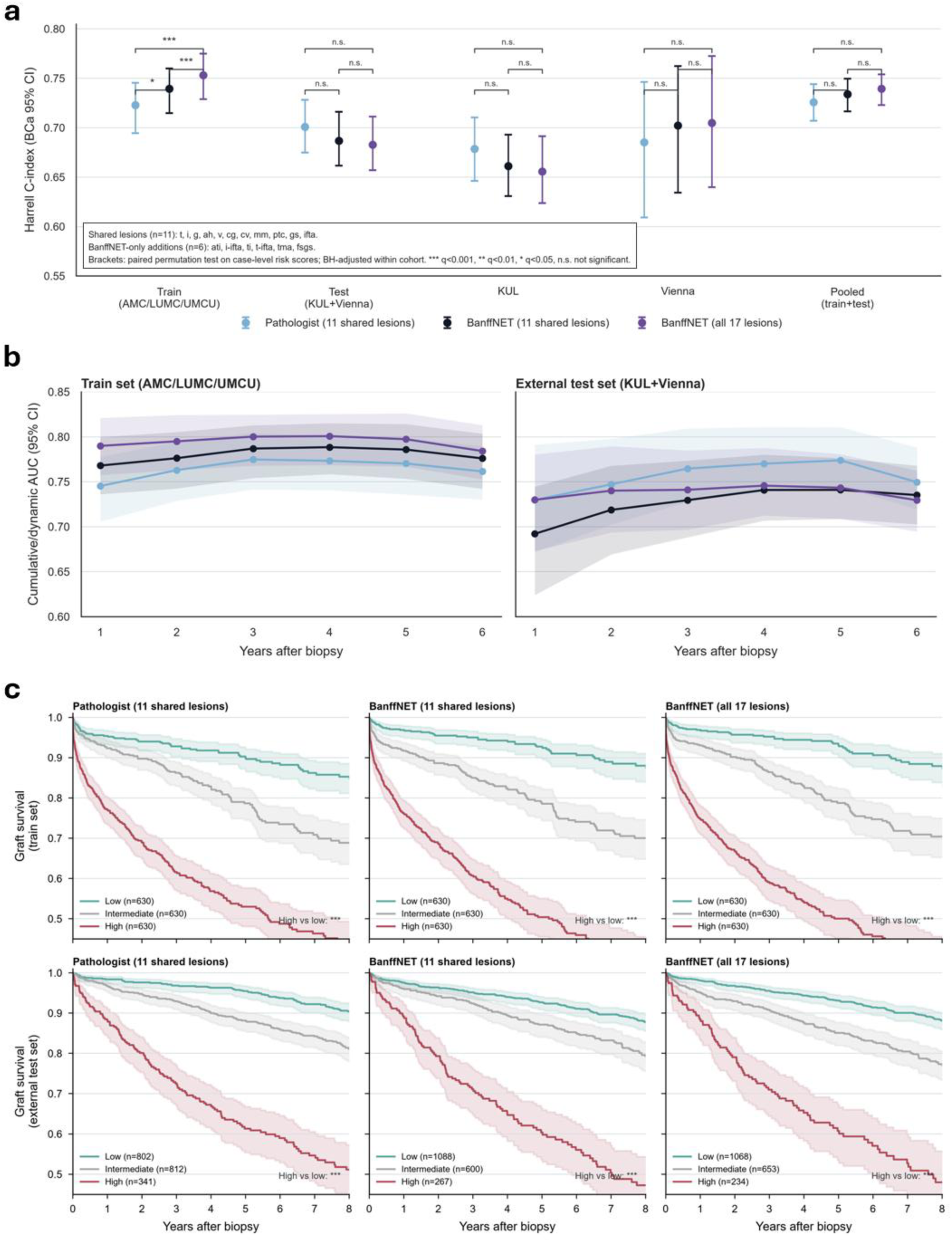
Comparison of BanffNET and pathologist Banff lesion scores for the association with death-censored graft failure. **a.** Overall discrimination of death-censored graft failure by multivariable models based on pathologist Banff scores, multivariable BanffNET models with matching 11 scores and the extended multivariable BanffNET model containing all 17 lesions. **b.** Cumulative-dynamic ROC curves for the discrimination of death-censored graft failure over time, t=0 is the time of biopsy. **c.** Patient stratification in risk tertiles (low, intermediate, high risk) according to multivariable pathologist Banff and BanffNET lesion score models shows comparable risk stratification between BanffNET and pathologist Banff score models. ***P<0.001, **P<0.01, *P<0.05, n.s., not significant.

### BanffNET scores align with molecular diagnostics and capture biologically relevant rejection processes

To assess the molecular and biological grounding of BanffNET predictions, we examined their alignment with multiple layers of biopsy-derived molecular data. In the Vienna cohort, where histological slides were linked to the clinically deployed Molecular Microscope Diagnostic System (MMDx) reports^16^, BanffNET lesion scores more closely aligned with MMDx-derived molecular probabilities of the same lesions than pathologist-assigned scores, particularly in the context of antibody-mediated rejection (AMR) (**Figure 5a-b; Extended Data Figure 12**).

**Figure 5.**
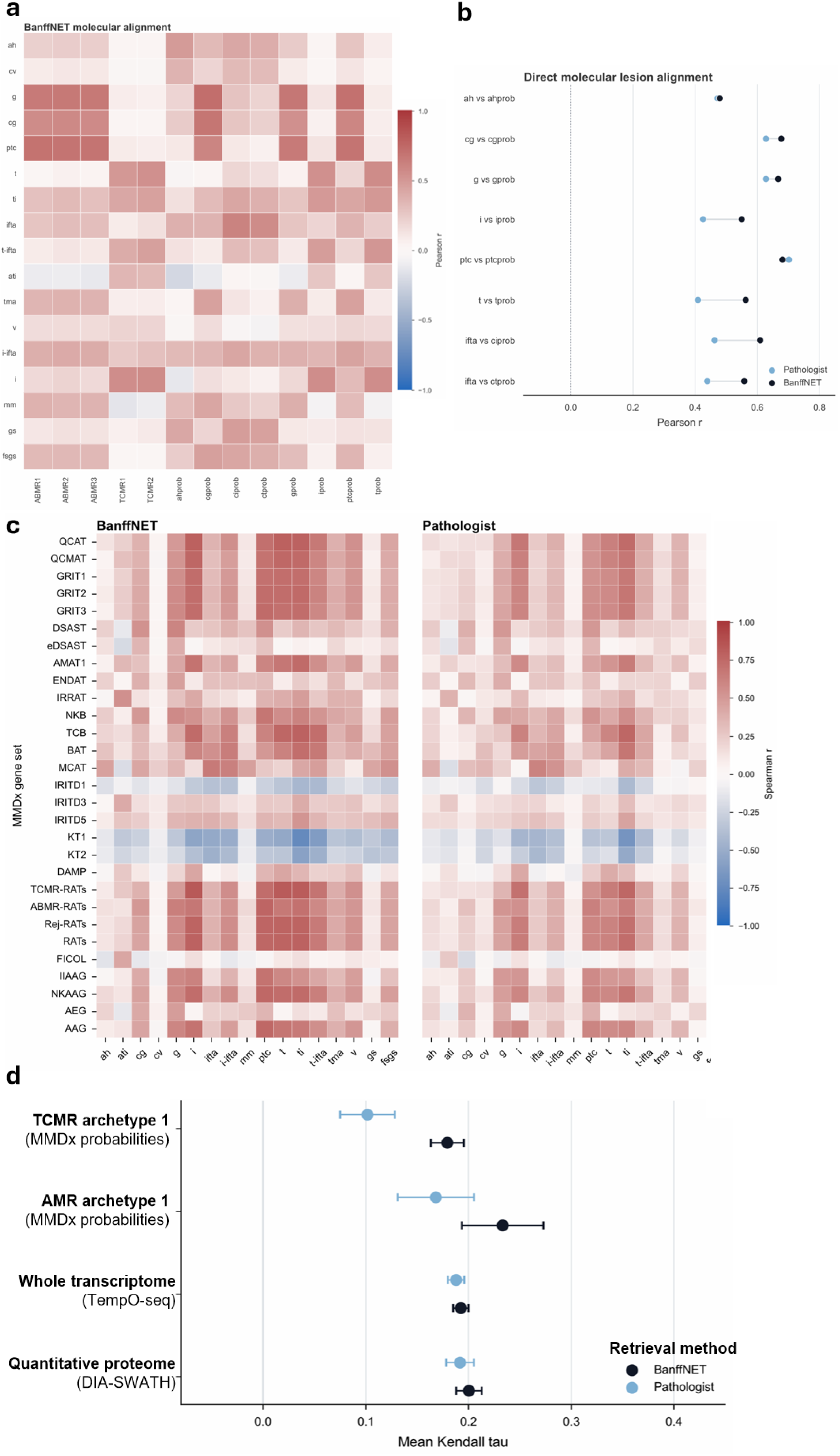
Alignment of BanffNET with molecular data. **a.** Correlation matrix showing strong correlations with molecular lesion-specific probabilities as well as rejection archetypes of AMR and TCMR. **b.** Difference in lesion-specific correlations with MMDx molecular lesion probabilities for BanffNET scores (black) versus pathologist Banff lesion scores (blue). **c.** Correlations of BanffNET and pathologist Banff lesion scores with MMDx pathogenesis-based transcript gene set scores derived from biopsy whole transcriptome analysis (TempO-seq). **d.** Molecular similarity search analysis for biopsies based on MMDx archetype probabilities (Vienna), across the full transcriptomic representation (targeted whole-transcriptome TempO-seq data, Leiden), and across the full quantitative proteomic representation (high-resolution data-independent acquisition data, Amsterdam), showing better retrieval of biopsies based on molecular content with BanffNET compared with pathologist Banff scores (a higher mean Kendall tau indicates greater similarity between biopsy rankings derived from the molecular and histological data representations across all biopsies).

To further probe this biological correspondence, we performed single-sample gene set enrichment analysis (ssGSEA) using previously defined transcriptional signatures (MMDx pathogenesis-based transcripts^16^) of alloimmune injury, including cytotoxic T cell activity, macrophage infiltration, interferon-γ signaling, endothelial damage, and tissue remodeling. BanffNET scores demonstrated stronger and broader enrichment across these rejection-associated pathways than pathologist Banff lesion scores (**Figure 5c**), underscoring their capacity to reflect immune and tissue-specific processes that underlie histological lesions.

Finally, we evaluated whether BanffNET could improve biopsy-level indexing based on molecular similarity (see **Online Methods**). In multiple content-based retrieval tasks (**Figure 5d**), BanffNET scores more accurately retrieved biopsies with similar molecular profiles than pathologist Banff scores. This included TCMR and AMR archetype probabilities derived from MMDx analysis (Vienna cohort), targeted whole-transcriptome profiles measured by TempO-seq (Leiden cohort, N=274), and high-resolution quantitative proteomic profiles measured by data-independent acquisition (Amsterdam cohort, N=276). These findings not only highlight BanffNET’s potential as a histological decision-support algorithm, but also as a bridge between digital pathology and molecular tissue profiling.

## Discussion

BanffNET establishes a unified, interpretable, and generalizable deep learning framework that overcomes the core technical limitations of current artificial intelligence applications in histology: lack of transparency, poor calibration, and reliance on discrete, threshold-based classification. By combining large-scale, self-supervised representation learning^8,17^ with domain-specific probabilistic graphical modeling, BanffNET introduces a novel Bayesian multiple-instance deep learning (MIL) architecture tailored to tissue pathology. Where current weakly supervised models are parameter-heavy and rely on often unstable attention mechanisms^9,18,19^, BanffNET embeds histological reasoning directly into its aggregation layer through lesion-specific probabilistic operators^20,21^: Noisy-OR Instance Pooling (NIP) for focal severity assessments and Gated Instance Pooling (GIP) for diffuse, proportion-based injury. This design enables the system to learn continuous, calibrated, and anatomically resolved lesion quantifications exclusively from slide-level supervision across multi-center datasets, despite varying staining protocols, and diverse scanner workflows.

A fundamental challenge in standardizing transplant pathology is that the established gold standard, human-assigned Banff lesion scores, is inherently subjective, semi-quantitative, and bounded by arbitrary ordinal cut-offs that fail to capture disease as a biological continuum. Because histological consensus is flawed by substantial interobserver variability^3^, benchmarking computational models against human annotations alone is insufficient to establish biological fidelity. Across an international reader study of 67 renal pathologists^11^, BanffNET matched or exceeded expert review across canonical lesions, outperforming individual human readers in evaluating subtle features with historically low rater reliability, such as peritubular capillaritis, interstitial inflammation, and tubulitis^11^. More importantly, when validated against objective, orthogonal anchors of graft integrity, BanffNET scores demonstrated comparable or superior clinical and biological alignment compared to pathologist assessments. Continuous model outputs aligned more closely with molecular representations of rejection, enriched more broadly for pathogenesis-based transcript signatures of alloimmune injury^16^, and provided comparable prognostic discrimination, with improvement in selected analyses. These findings indicate that probabilistic lesion modeling captures sub-threshold tissue injury that is insufficiently captured by ordinal human grading, offering a reliable, and by design reproducible substrate for refining diagnostic criteria and risk thresholds in transplantation, in turn further improving the current Banff classification system^22^.

Methodologically, BanffNET represents a significant conceptual advance over existing state-of-the-art weakly supervised learning architectures in computational pathology. Compared to high-capacity attention-based MIL frameworks (such as ABMIL^9^ and TransMIL^18^), BanffNET’s low-capacity NIP and GIP Bayesian regression heads^23^ achieve superior predictive accuracy while reducing parameter counts by more than 2,500-fold, offering a highly computationally efficient solution that processes gigapixel whole-slide images in seconds^24^. Beyond computational efficiency, the probabilistic design of NIP and GIP overcomes the visual ambiguity of standard attention heatmaps^25^. When coupled with Generic Attention Explainability (GAE)^14^, BanffNET generates localized, pixel-level probability maps that align with underlying anatomical structures, mapping leukocyte infiltration inside microvessels or early basement membrane duplication with a granularity that surpasses diffuse attention-based localization. Furthermore, despite training each lesion model independently without explicit multi-task regularization, BanffNET preserved reproducible patterns of lesion co-occurrence across independent test cohorts with higher matrix consistency than human scoring, suggesting that its outputs capture relationships consistent with known pathophysiological patterns of graft injury.

From a clinical and translational perspective, BanffNET addresses the growing crisis of diagnostic variability and severe global workforce shortages in specialized renal pathology services (86% of countries report a shortage of renal pathologists)^26,27^. In routine deployment, the framework can serve as an accessible decision-support system for generalist or junior pathologists as well as nephrologists, standardizing kidney graft biopsy interpretation and flagging subtle, multifactorial lesions for expert review. Moreover, BanffNET demonstrates unique utility as a bridge between digital morphology and molecular profiling. In content-based retrieval tasks across independent cohorts and molecular modalities, BanffNET lesion scores outperformed expert annotations in indexing and retrieving biopsies with similar whole-transcriptome and proteomic profiles. This capacity to identify histological patterns associated with molecularly conserved phenotypes from routine, inexpensive histological stains positions BanffNET as a potentially valuable tool for enriching clinical trials and large observational studies, guiding targeted molecular testing, and enabling multi-modal precision diagnostics in resource-constrained environments.

The modular architecture underlying BanffNET extends beyond kidney transplantation, providing a transferable blueprint for complex, subjective, and spatially heterogeneous diagnostic systems across computational pathology. In oncology, major tumor grading frameworks, such as Gleason scoring in prostate cancer, Nottingham grading in breast cancer, or histological subtyping in various tumors, all suffer from interobserver variability and spatial heterogeneity that parallel the challenges of the Banff classification. Implementing lesion-specific probabilistic pooling could similarly convert these discrete, ordinal grading systems into continuous, prognostic risk spectra. Likewise, in chronic inflammatory and autoimmune diseases such as lupus nephritis, inflammatory bowel disease, or metabolic liver disease, where overlapping focal and diffuse processes dictate therapeutic management, the dual NIP/GIP architecture offers a principled methodology for disentangling coexisting histological features with high precision under weak supervision.

Several limitations of this study warrant consideration. First, because BanffNET was trained using pathologist-assigned slide labels, the model is inherently anchored to an imperfect histological gold standard that may not encompass all biologically relevant tissue manifestations. Second, our training and validation cohorts were primarily derived from European academic medical centers. Validation in more ethnically and geographically diverse patient populations is required to ensure fair and unbiased clinical performance. Third, the current iteration of the BanffNET system uses a general-purpose (pre-trained) histology foundation model with limited training on kidney biopsy data, which could affect the quality of the extracted features. Finally, translating these probabilistic scores into actionable clinical workflows will require formal integration into the international Banff rejection classification system, alongside prospective randomized clinical trials to evaluate its real-world impact on diagnostic turnaround times, treatment decision-making, and rejection-specific graft survival risk stratification.

In summary, BanffNET advances computational pathology from a paradigm of discrete, opaque classification toward an interpretable, probabilistic quantification of tissue injury. By bridging the gap between digital morphology, molecular phenotypes, and clinical outcomes, this framework provides a scalable foundation for standardized and biologically grounded diagnostic precision in transplantation and beyond.

## Online Methods

### Detailed description of the cohorts and data curation

#### Amsterdam University Medical Center, Amsterdam, Netherlands

Amsterdam UMC (abbreviated as AUMC) is a tertiary university hospital affiliated to the University of Amsterdam and the VU University Amsterdam covering nephrology care for the northwestern region of the Netherlands plus a transatlantic transplantation program for multiple Caribbean islands within the Kingdom of the Netherlands (Aruba, Bonaire, Curacao, Saba, Sint Eustatius, Sint Maarten). The transplant population of Amsterdam UMC is the most ethnically diverse case mix in the Netherlands and includes patients from Surinamese, Afro-Caribbean, Turkish, Moroccan and Ghanaian descent. The Netherlands is part of the Eurotransplant program. Patients who underwent a kidney biopsy between 2000 and 2019 were included in the study (preimplantation biopsies excluded). Local data collection was approved by the institutional review board under registration number 19.260 and informed consent was waived. All biopsies were scanned with a Philips Ultrafast whole slide image (WSI) scanner at a resolution of 0.25 micrometer per pixel. All cases were scored by three pathologists blinded to the clinical diagnosis and from each other’s scores. A total of 1048 biopsies (3133 WSIs) were included in the BanffNET **training** cohort. On 276 biopsies from the cohort (as described by Hofstraat et al.^28^), data-independent acquisition (DIA) mass spectrometry (LC-MS/MS) was performed on the same formalin-fixed paraffin-embedded (FFPE) tissue block as the WSI, resulting in quantification of >5.000 proteins per sample. Standard nearest-neighbor imputation and principal component dimensionality reduction were applied.

### Leiden University Medical Center, Leiden, Netherlands

Leiden University Medical Center (abbreviated as LUMC) is a tertiary university hospital affiliated to Leiden University covering nephrology care for the western Randstad region of the Netherlands. As a Center of Expertise for rare kidney diseases, the transplant population is enriched with patients who suffer from systemic autoinflammatory and autoimmune disorders, including lupus nephritis, antiphospholipid syndromes, vasculitis, and both genetic and acquired complement disorders. The Netherlands is part of the Eurotransplant program. Adult patients who underwent a kidney biopsy between 2011 and 2024 were included in the study (preimplantation biopsies excluded). Local data collection was approved by the institutional review board under registration number W2020.031 and informed consent was waived. All biopsies were scanned with a Philips Ultrafast WSI scanner at a resolution of 0.25 micrometer per pixel. A total of 913 biopsies (2659 WSIs) were included as BanffNET **training** cohort. On 274 biopsies from the LUMC cohort, whole transcriptome targeted RNA sequencing with Templated Oligo-sequencing (TempO-Seq) was performed. Probe sequences were first reconciled with the most recent NCBI gene symbol annotation (release 2024-12-10). Probes exhibiting <10 counts per million (CPM) in >95% of samples were discarded, leaving 14.220 high-confidence probes. Counts from probes mapping to the same gene were subsequently aggregated to obtain gene-level expression values (Ngenes = 12.740). Samples with sub-threshold sequencing depth (library size = mean – 2*SD; 3.459.619 reads) or mapping efficiency (percentage mapped = mean – 2*SD; 38%) were excluded (n = 36). Batch effects arising from the five sequencing runs were mitigated with ComBat-seq (sva v3.56.0, Bioconductor).

### University Medical Center Utrecht, Utrecht, Netherlands

UMC Utrecht (abbreviated as UMCU) is a tertiary university hospital affiliated to Utrecht University covering nephrology care for the central region of the Netherlands. The transplant population of UMCU consists mainly of patients from Dutch descent. The Netherlands is part of the Eurotransplant program. Adult patients who underwent a kidney biopsy between 2000 and 2019 were included in the study (preimplantation biopsies excluded). Local data collection was approved by the institutional review board under registration number 19.482 and informed consent was waived. All archival biopsies were scanned retrospectively with a Philips Ultrafast WSI scanner from 2000-2016 and with a Hamamatsu XR scanner as part of the daily clinical workflow from 2016-2019. All WSIs were scanned at 0.25 micrometer per pixel resolution. A total of 583 biopsies (1741 WSIs) were included in the BanffNET **training** cohort.

### University Clinic RWTH Aachen, Aachen, Germany

University Clinic RWTH Aachen (abbreviated as UKA) is a tertiary university hospital covering nephrology care for the western region of Germany including a longstanding kidney transplantation program. The transplant population is mostly Caucasian but also includes other ethnically diverse groups such as patients from Turkey or India. Germany is part of the Eurotransplant program. Factually anonymized data from patients who underwent a kidney biopsy between 2017 and 2021 were included in the study (preimplantation biopsies excluded). Local data collection was approved by the institutional review board under registration number EK 23-094 and informed consent was waived. All biopsies were scanned with a Aperio AT2 WSI scanner with a resolution of 0.25 micrometer per pixel. A total of 95 biopsies (285 WSIs) were included as part of the BanffNET **validation** cohort.

### University Hospitals Leuven, Leuven, Belgium

UZ Leuven is a tertiary university hospital affiliated to KU Leuven University (abbreviated as KUL), the largest university hospital in Belgium. In 2024, 387 organs were transplanted at UZ Leuven, or 32,7% of all transplanted organs in Belgium. This makes UZ Leuven also the largest of the 8 transplant centres in Belgium. Belgium is part of the Eurotransplant program. Adult patients who underwent a kidney transplant biopsy between March 2004 and March 2020 were included in the study (preimplantation biopsies excluded). Local data collection was approved by the institutional review board under registration number S64006 (clinicaltrials.gov NCT06505200) and informed consent was waived. All biopsies were scanned with a Philips Ultrafast WSI scanner at a resolution of 0.25 micrometer per pixel. A total of 3696 biopsies (9708 WSIs) were included in the BanffNET **validation** cohort.

### Medical University Vienna and AKH Wien, Vienna, Austria

The Medical University of Vienna (abbreviated as VIENNA), together with its clinical partner AKH Wien (General Hospital of Vienna), forms one of the largest academic medical centers in Europe. As a tertiary care university hospital, AKH Wien provides comprehensive nephrology services for the eastern region of Austria, with a particular emphasis on complex renal pathologies and transplant nephrology. The transplant program at MedUni Vienna/AKH includes national and international referrals, with a diverse patient population encompassing Central and Eastern European as well as Middle Eastern descent. Austria is part of the Eurotransplant program. Patients who underwent a kidney transplant biopsy between 2013 and 2019 were included in the study (preimplantation biopsies excluded). Local data collection was approved by the institutional ethics committee under registration number 1600/2020, and informed consent was waived. All biopsies were scanned with a 3DHISTECH Pannoramic 250 III Flash scanner for the acquisition of WSIs at a resolution of 0.25 micrometer per pixel. A total of 222 biopsies (657 WSIs) were included as part of the BanffNET **validation** cohort. For each biopsy, a 3-mm core segment was placed in RNAlater and shipped at room temperature to the Alberta Transplant Applied Genomics Centre for microarray analysis via the Molecular Microscope Diagnostic System (MMDx). Gene expression was measured using GeneChip PrimeView U219 arrays and processed in R with BioBase v.2.64.0. Data were normalized to the K1208 kidney allograft biopsy reference set using robust multiarray averaging, enabling MMDx classifier score prediction. Machine-learning-derived ensemble classifiers were previously developed using kidney transplant or murine biopsy data^16^. For each biopsy, molecular probability scores and rejection archetype scores were calculated as described by Halloran and colleagues^16^.

### University Hospital of Parma, Parma, Italy

The University Hospital of Parma (abbreviated as PARMA) is a medium-to-large tertiary care center located in Northern Italy. The transplant population consists of adult patients from across the country, primarily of Italian descent, including those from expanded criteria donors. A protocol kidney biopsy at four months after transplantation is part of the standardized follow-up schedule. Local data collection was approved by the institutional review board under registration number 0046283, and informed consent was waived. Allograft kidney biopsies, either protocol or indication biopsies, performed between January 2021 and August 2024, for which scanned images were available, were included in the study. All WSIs were acquired using the Motic Easy Scanner at a resolution of 0.13 μm/pixel (80x) and stored in SVS format. Corresponding Banff scores were assigned by pathologists at the time of diagnosis and collected from pathology reports. A total of 414 biopsies (1214 WSIs) were included as part of the BanffNET **validation** cohort.

### Radboud University Medical Center Nijmegen, Nijmegen, Netherlands

Radboud University Medical Center Nijmegen (abbreviated as RUNMC) is the teaching hospital affiliated with the Radboud University Nijmegen, in the city of Nijmegen. It is one of the largest and leading hospitals of The Netherlands, providing supraregional tertiary care for residents of a large part of the eastern section of The Netherlands. RUNMC is an international referral center for rare renal disorders and complement-mediated diseases. It is one of the seven university medical centers that perform kidney transplantation, conducting roughly 125 to 150 procedures annually. Approximately 80 of these transplantations include living donors. Local data collection was approved by the institutional review board under registration number 2022-13686 and informed consent was waived. All biopsies were digitized using a Pannoramic P1000 whole slide scanner (3DHISTECH, Hungary) at a resolution of 0.24 µm per pixel. A total of 282 biopsies (823 WSIs) from 186 patients taken for cause during 2016-2017 period were included in the BanffNET **validation** cohort.

### LUMC global reader study (reader study)

The LUMC global reader study^11^ is a fully online digital reader study that included 36 cases (108 WSIs; an HCE, PAS and silver staining for each case), not overlapping with the LUMC training cohort. Slides were prospectively scanned with the Philips Ultrafast whole slide image scanner at a resolution of 0.25 micrometer per pixel for primary clinical diagnostics. Digitized WSIs were uploaded to the online platform Slidescore (https://www.slidescore.com) and fully anonymized. Included were a total of 67 pathologists from 50 cities in 24 countries: Argentina (2), Australia (4), Austria (3), Bangladesh (2), Belgium (1), Brazil (1), Canada (4), Egypt (1), France (4), Germany (2), India (3), Indonesia (1), Iran (1), Italy (6), Kuwait (1), Nigeria (1), Saudi Arabia (1), South Africa (1), Sweden (1), Taiwan (1), Thailand (2), The Netherlands (3), United Kingdom (4), United States of America (17). The study spans all permanently inhabited continents (Europe, Americas, Africa, Asia, Australia) and 13 of 22 (59%) geographical subregions as defined by the UNSD. All participants are renal pathologists experienced with scoring the Banff classification system with an average of 10 years (range 1 - 30 years) of experience. For each of the 36 cases, a single integrated electronic case report form (eCRF) was created in Slidescore and cases were presented to the pathologists in a per-person randomized order. Pathologists were asked to assess the following scores: total number of glomeruli, total number of globally sclerosed glomeruli, g (0-3), cg (0-3), mm (0-3), i (0-3), t (0-3), ptc (0-3), v (0-3 or absent), ti (0-100% with 10% increments), ifta (0-100% with 10% increments), i-ifta (0-3), t-ifta (0-3), cv (0-3 or absent), ah (0-3), acute tubular injury (present/absent), thrombotic microangiopathy (present/absent), and focal and segmental glomerulosclerosis (present/absent). The LUMC global reader study is used as a **validation** cohort.

### Data to train focal and segmental glomerulosclerosis and global glomerulosclerosis

The percentage of glomeruli affected by global glomerulosclerosis (gs) is structurally reported in native and transplant kidney disease. For focal and segmental glomerulosclerosis (fsgs), quantification of the affected glomeruli is not consistently performed and is often reported as present or absent. We curated a fully anonymized collection of a total of 31972 WSIs from 11266 biopsies across all available native and transplant cases to train, validate and test both algorithms. A detailed breakdown of the number of WSIs/biopsies that were used to train, validate and test gs and fsgs can be found in **Extended Data Table 18**. As fsgs and gs are not part of the Banff system, we report them as extra BanffNET lesion scores, but did perform an extensive head-to-head comparison with the pathologist scores. Similar to the BanffNET lesion scores, separate models were trained for the HCE, PAS and Silver staining.

### Digital tissue processing and feature extraction

WSIs were analysed using a standardized pipeline to generate image patches suitable for downstream feature extraction. Regions of interest (ROIs) were manually annotated by expert pathologists to exclude control tissue and focus the analysis on diagnostically relevant areas. To ensure that extracted patches contained tissue, a binary mask was first generated using a deep segmentation model. A diverse set of 726 biopsy WSIs from multiple institutions stained with hematoxylin and eosin (HCE), Jones’ silver (Silver) or periodic acid-Schiff (PAS), containing a large variation of background artifacts, were annotated for their biopsy contours. A UNet++ architecture^29^ with a MobileNetV2 backbone^30^ was trained and an additional 49 ROIs were used for validation. The segmentation model was initialized with ImageNet-pretrained weights and optimized using the Adam algorithm with weight decay until convergence on the validation set. The segmentation model was optimized using a composite Dice-Binary Cross-Entropy (Dice-BCE) loss function with label smoothing to ensure robust tissue boundary detection. During training of the segmentation model, a comprehensive augmentation strategy was applied to enhance model generalizability across stain types and acquisition conditions. Augmentations included geometric transformations (e.g., random resized cropping, flipping, rotation, and perspective distortions), color perturbations (e.g., CLAHE, brightness/contrast shifts, gamma adjustments, and hue-saturation-value modifications), blurring and noise injection (e.g., Gaussian and motion blur, and Gaussian noise), as well as structural distortions (e.g., elastic, grid, and optical distortions). We applied CoarseDropout to simulate occlusions and artifacts. At inference time, input regions of interest were downsampled for rapid tissue segmentation prior to patch extraction. Contrast-limited adaptive histogram equalization (CLAHE) was applied to the image. A morphological closing operation with a kernel size of 7 pixels was then applied to the mask to eliminate small holes and improve tissue continuity.

The resulting binary segmentation masks were used to guide patch extraction from whole-slide images (WSIs) at a fixed resolution of 0.5 µm/pixel. Each biopsy *b_i_* i is cut into patches *P_i_* = {*p_ij_*} for *j* = 1,2, …, *s_i_*, where *s_i_* denotes the number of patches. To reduce colour variability introduced by differing staining procedures, we applied Reinhard normalization to each patch in the biopsy. Because each staining type (e.g., HE, PAS, Silver) possesses distinct colour characteristics, we computed separate reference statistics in the LAB colour space for each staining type. These stain-specific reference values were then used to normalize each patch, yielding p*_ij_*. Patches were extracted in a non-overlapping manner, each 256×256 in pixel size. Patches containing sufficient tissue coverage, as determined by the binary mask, were retained for downstream processing. There were no restrictions on the size of the biopsy for training or testing as they could also be used for Banff scoring by the pathologists. There was no artifact filter as we believe that models should be robust and ignore tissue processing or scanning abnormalities during training and inference. Extracted image patches were resized to 224×224 pixels prior to feature extraction using the UNI histology foundation model^12^. Each normalized patch p*_ij_* is then represented by a feature vector extracted using UNI foundation model *f_UNI_* : *R^W^*^×*H*×3^ → *R^M^*, yielding.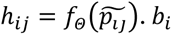 *b_i_* is then described by *H* = [ℎ_i1_,ℎ_i2_,…,ℎ_isi_]^T^ ∈ *R^si^*^×*M*^. Feature vectors were stored in HDF5 format (H5), with corresponding spatial coordinates preserved to allow mapping back to original WSI locations.

### Design and training of BanffNET

Given lesion scores *Y* = {*y*_1_, *y*_2_, …, *y_N_*} mapped to probabilities via min-max normalization, our goal is to model the probability that a biopsy *b_i_* exhibits a particular lesion *P*(*b_i_*) = *g_Θ_*(*H_i_*), *w*ℎ*ere g_Θ_*: *R^K^*^×*M*^ → [0,1] as the aggregation function parameterized by *Θ*. The following two aggregation functions were designed:

The Gated Instance Pooling (GIP), which computes:

1. 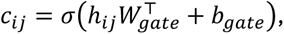
2. 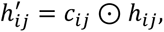
3. 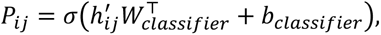,
4. 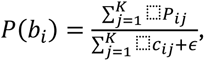, where *ε* is a small constant added for numerical stability.

The Noisy-Or Instance Pooling (NIP), which computes 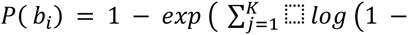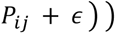.

Models were optimized using the Adam optimizer with a composite learning rate schedule incorporating warm-up and cosine annealing^31^. Training was performed using binary cross-entropy loss with a class-weighted loss. Evaluation metrics included area under the ROC curve (ROC-AUC), F1 score, balanced accuracy, and Pearson correlation. Feature bags of varying sequence lengths were padded and masked during batching to ensure correct loss computation. TensorBoard was used for real-time monitoring of performance metrics, confusion matrices, and runtime statistics. Model outputs were calibrated using standard post-hoc logistic scaling to yield slide-level severity scores. ABMIL^32^ and TransMIL^18^ models were trained using the same pipeline as NIP and GIP, with two key differences. First, to remain faithful to the original implementations, training was performed with a batch size of 1 (one bag at a time). Second, the learning rate schedule was adapted to improve convergence, with the following hyperparameters: an initial learning rate of 1 × 10^-5^, a final learning rate after warm-up of 1 × 10^-3^, and cosine annealing ranging between 1 × 10^-4^ and 1 × 10^-3^. All other training settings, including loss function, instance-level weighting, evaluation metrics, and feature preloading strategy, remained consistent with those used for NIP and GIP.

### Pixel-level lesion visualization

We generated relevance maps using the Generic Attention Explainability (GAE) method^14^, following Chefer and colleagues. This was applied to the full architecture consisting of the frozen UNI ViT backbone^8^ and the BanffNET lesion heads. For both the NIP variant (single linear layer) and the GIP variant (classification branch), the sigmoid activation was removed to ensure gradient stability in single-precision computations. During the backward pass, we intercepted the raw self-attention matrix A from each transformer block and computed the element-wise product with its class-specific gradient (∂log(p)/∂A). Relevance propagation maps were derived from transformer self-attention weights to localize structural features. Patches were densely sampled across the slide, and relevance maps were thresholded to highlight high-confidence lesion regions. The resulting attention maps were overlaid on the ROI with a transparency of 0.3.

### Content-based whole slide image retrieval with biopsy transcriptome or proteome as a gold standard

We assessed the alignment between Banff score–based and omics-based descriptors of kidney transplant biopsies using a similarity-based ranking procedure. For each biopsy *b_i_*, we defined two representations: (1) a Banff score vector 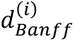, either assigned by a pathologist (or consensus by multiple pathologists) or calculated by BanffNET and (2) an omics descriptor 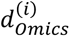 derived from proteomics or transcriptomics of the same kidney transplant biopsy. Missing omics values were imputed using k-nearest neighbors. To quantify pairwise similarity between biopsies *b_i_* and *b_j_*, we used Euclidean distance for the Banff vectors 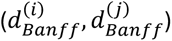, with smaller values indicating greater similarity, and cosine similarity for the omics descriptors 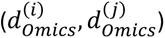, where higher values indicate greater similarity. Each biopsy *b_i_* thus induces two separate rankings of all other biopsies: one based on Banff scores (pathologist Banff versus BanffNET scores), 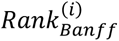, and one based on omics data (proteomics or transcriptomics), 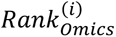 . We measured the concordance of these rankings for each biopsy *b_i_* using Kendall’s tau (*τ*^(*i*)^)^33^, and then averaged across all biopsies to obtain 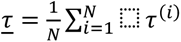. A higher *τ* indicates stronger agreement between Banff score- and omics-based similarity measures, implying that the Banff scores capture biologically meaningful patterns reflected in the omics data. We repeated this analysis separately using pathologist-assigned and model-predicted Banff scores and compared the resulting *<u>τ</u>*.

### Statistical analysis

A stratified train–validation split was used for every lesion and stain model. A class-balanced validation hold-out set was created across ordinal classes for early stopping and checkpoint selection. In the training cohort, associations between continuous BanffNET scores and pathologist Banff scores were evaluated using Mann-Whitney U tests (for binary pathologist scores), Jonckheere-Terpstra ordered-trend tests (for ordinal pathologist scores), and Spearman’s rank correlation tests (for continuous pathologist scores). In the validation cohort, we performed the analysis for binary pathologist scores stratified by external cohort using the Van Elteren test. For ordinal and continuous pathologist scores, effect sizes were quantified using Spearman’s rho and pooled via a random-effects meta-analysis as follows: 1) calculate the Spearman rho for each cohort, 2) apply Fisher’s z-transformation to each correlation coefficient using Spearman-adjusted sampling variances, 3) estimate between-center heterogeneity by fitting a random-effects model with the non-parametric Paule-Mandel estimator, 4) pool the effect sizes using the Hartung-Knapp-Sidik-Jonkman (HKSJ) method, and 5) convert the pooled effect size estimate and the upper and lower bounds of the 95% HKSJ confidence interval back to the original correlation scale. Cross-cohort similarity of lesion– lesion correlation matrices were tested with the Mantel statistic^34^ (10,000 permutations). An elastic-net penalized Cox model, implemented in *scikit-survival*, with default hyperparameters (l1_ratio = 0.5; 100 alphas, alpha_min_ratio=’auto’) predicted death-censored graft failure. Follow-up was truncated at six years and observations beyond this horizon were right-censored. The lesion covariates were entered either as BanffNET lesion scores or as pathologist lesion scores rescaled to 0-1. Models were trained on biopsies from Amsterdam, Leiden and Utrecht and evaluated on biopsies from Leuven and Vienna for which graft survival data was available. Discrimination was assessed with Harrell’s C-index^35^, and time-dependent AUCs (Δt = 30 days) with 95 % confidence bands from 1000 patient-level bootstrap resamples; differences between models based on BanffNET scores and pathologist scores were tested on the same bootstrap distribution. Akaike Information Criterion (AIC) was used to test model fit and parsimony. At biopsy time, log-transformed serum creatinine and UPCR were related to individual lesion scores using ordinary least-squares (OLS) univariable and multivariable models; β estimates are reported with 95 % confidence intervals and two-sided p-values. Single-sample GSEA (ssGSEA)^36^ was applied to log₂-transformed counts per million (log₂-CPM) TempO-seq data obtained from the same biopsy block using the MMDx pathogenesis-based transcript gene-set collection^16^. For each lesion, Spearman’s rank correlation coefficients with ssGSEA scores were computed separately for BanffNET lesion scores and pathologist lesion scores.

### Computing hardware and software

All models were trained on an HPC node (Intel Xeon Gold 6234, 256 GB RAM, dual NVIDIA RTX 6000 GPUs, CentOS 8). The ROI binarization and BanffNET models were developed using Python 3.10 and PyTorch 1.13.1 with architectures from Segmentation Models PyTorch (v0.3.3). Data augmentation utilized Albumentations (v1.4.11), with image processing via OpenCV (v4.10.0) and scikit-image (v0.24.0). GPU training leveraged CUDA 11.6 and TensorBoard (v2.15.1) logging. Feature extraction and model inference were implemented in PyTorch utilizing GPU acceleration for efficient computation; model configurations used HuggingFace transformers (v4.36.2) and timm (v1.0.3). Statistical analyses, including correlation analysis, Mantel tests (custom implementation via SciPy v1.11.4), survival modeling (scikit-survival v0.23.1), were conducted in Python 3.12 (managed via Conda), with pandas (v2.2.2) supporting correlation computations. Visualizations utilized matplotlib (v3.9.1) and seaborn (v0.13.2). Gene Set Enrichment Analysis (GSEA) was performed in R (v4.4.1) using GSVA (2.0.4), GSEABase (1.68.0), clusterProfiler (4.14.4), fgsea (1.32.2), ComplexHeatmap (2.22.0), and circlize (0.4.16).

### Ethics approval

The Institutional Review Board (IRB) of the Amsterdam UMC approved the retrospective design of the study under number 19.260. Local IRB approval for data use can be found under the section “Detailed description of the cohorts and data curation”.

### Reporting summary

TRIPOD+AI and CLAIM reporting guidelines are available as online content linked to this article^37,38^.

## Supporting information

CLAIM checklist

TRIPOD+AI checklist

## Acknowledgements

This project has received funding from the European Union’s Horizon Europe research and innovation programme under grant agreement N° 101072891 and the Dutch Kidney Foundation grant N° 17OKG23 (JK). PB is supported by the German Research Foundation (DFG, Project IDs 322900939 C 445703531 C 552234081), European Research Council (ERC Consolidator Grant No 101001791), Federal Ministry of Research, Technology, and Space (BMFTR, 01KX2524), and the Innovation Fund of the Federal Joint Committee (Transplant.KI, No. 01VSF21048). During the preparation of this work, the authors used Gemini and Claude to assist with rewriting the manuscript text and schematic visualizations. All outputs created by generative artificial intelligence were validated by the authors.

## Data availability

Data in this study contain privacy-sensitive clinical information and cannot be shared online. Access to data will require approval of a study protocol and a data sharing agreement with each of the individual university medical centers involved in the study. All requests regarding data availability and legal procedures can be addressed to the principal investigator of this study (JK) at.

## Code availability

Code will be made available upon publication.

## Author contributions

GB (study lead) and JK (principal investigator) designed the study and were responsible for study oversight; CP, JH, RHB, GEB, TQN, SF and JK were involved in train data collection, data curation and/or labeling; AT, DvM, RB, DLH, ASM, MK, NK, GB, PFH, DvdH, SM, JHV, SH, LBH, EJS, ADvZ, ASN, FJB, IBB, GC, BvdW, TTP, GMR, EF, UM, JJTH, FT, FF, AAA, MD, GLC, PK, PB, MN, APJdV and JK curated and provided validation data; 67 pathologists (LGRS) scored the biopsies in the reader study; GB trained the models and performed all analyses with input from JK; JD, YKOT, HPS, MN, APJdV and JK were (in part) involved in supervision of the work; GB and JK wrote the manuscript; GB, MN, APJdV and JK provided critical review of the manuscript;. All authors reviewed the content of the manuscript prior to submission. No patient or public consultation took place during the development of BanffNET.

---

**Extended Data Figure 1.**
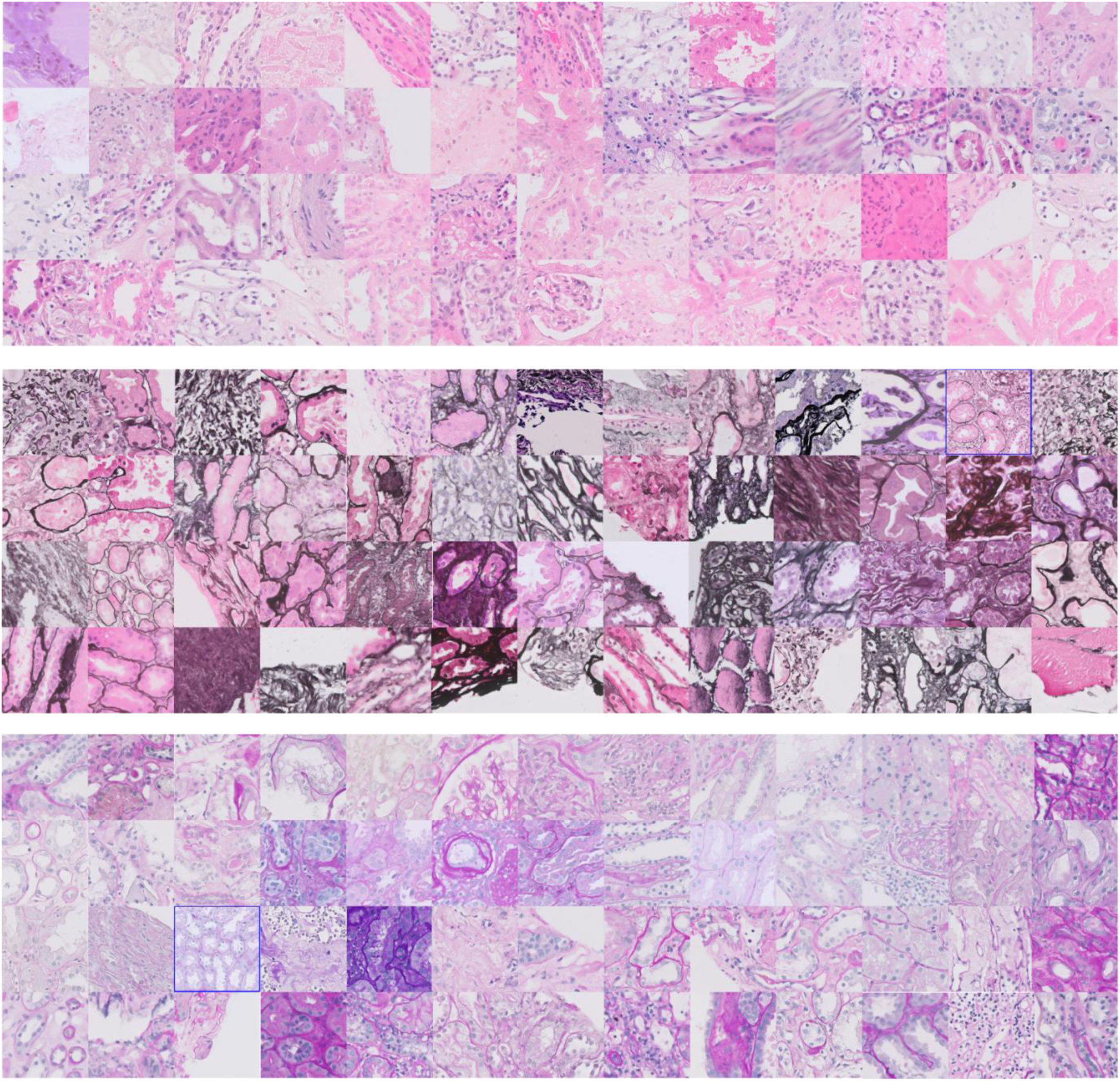
Mosaic of randomly sampled patches from the train cohorts. Patches with dimensions of 256 × 256 pixels were extracted at 0.5 micrometer per pixel resolution. Upper panel = HCE, middle panel = silver and lower panel = PAS.

**Extended Data Figure 2.**
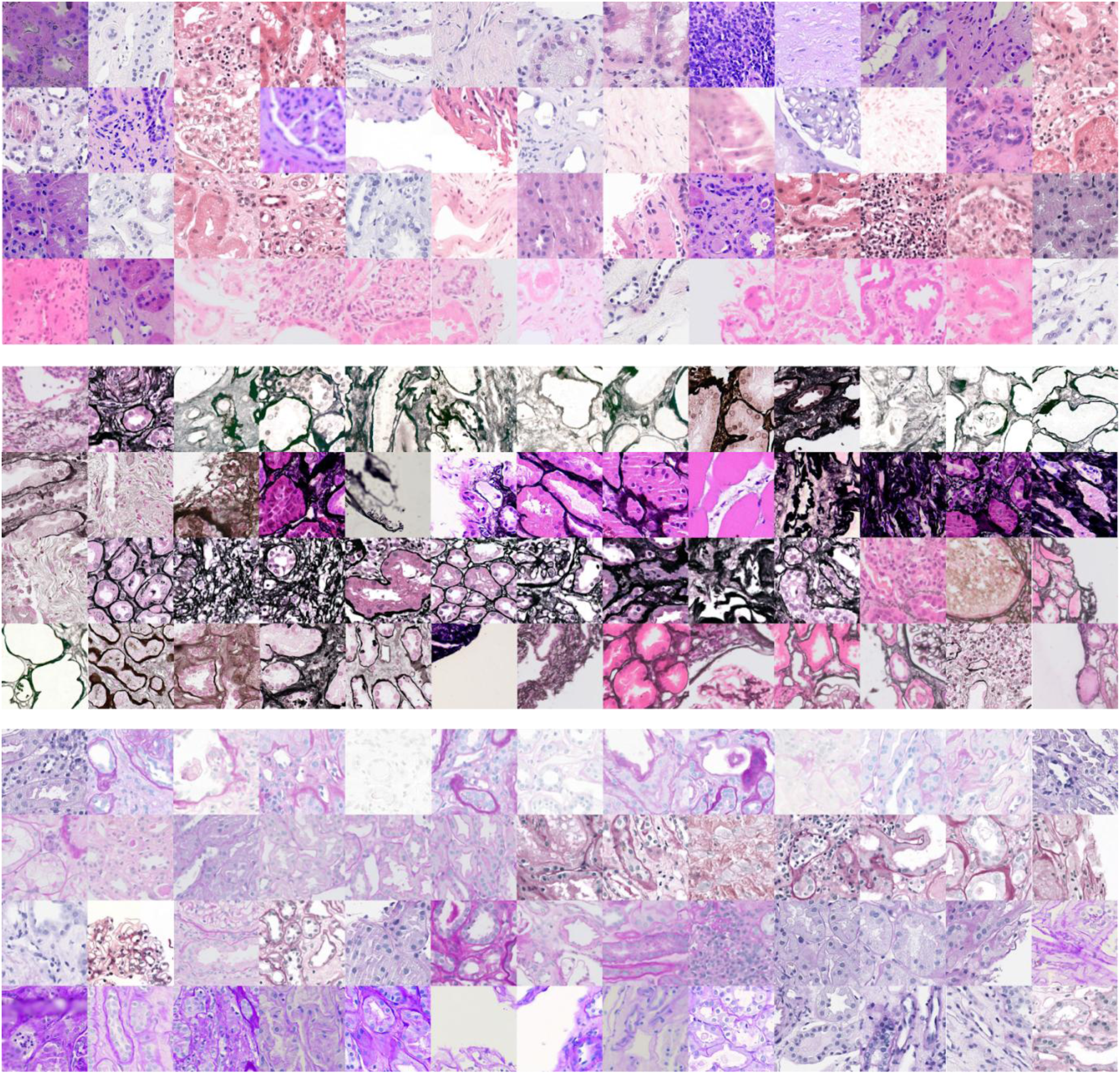
Mosaic of randomly sampled patches from the validation cohorts. Patches with dimensions of 256 × 256 pixels were extracted at 0.5 micrometer per pixel resolution. Upper panel = HCE, middle panel = silver and lower panel = PAS.

**Extended Data Figure 3.**
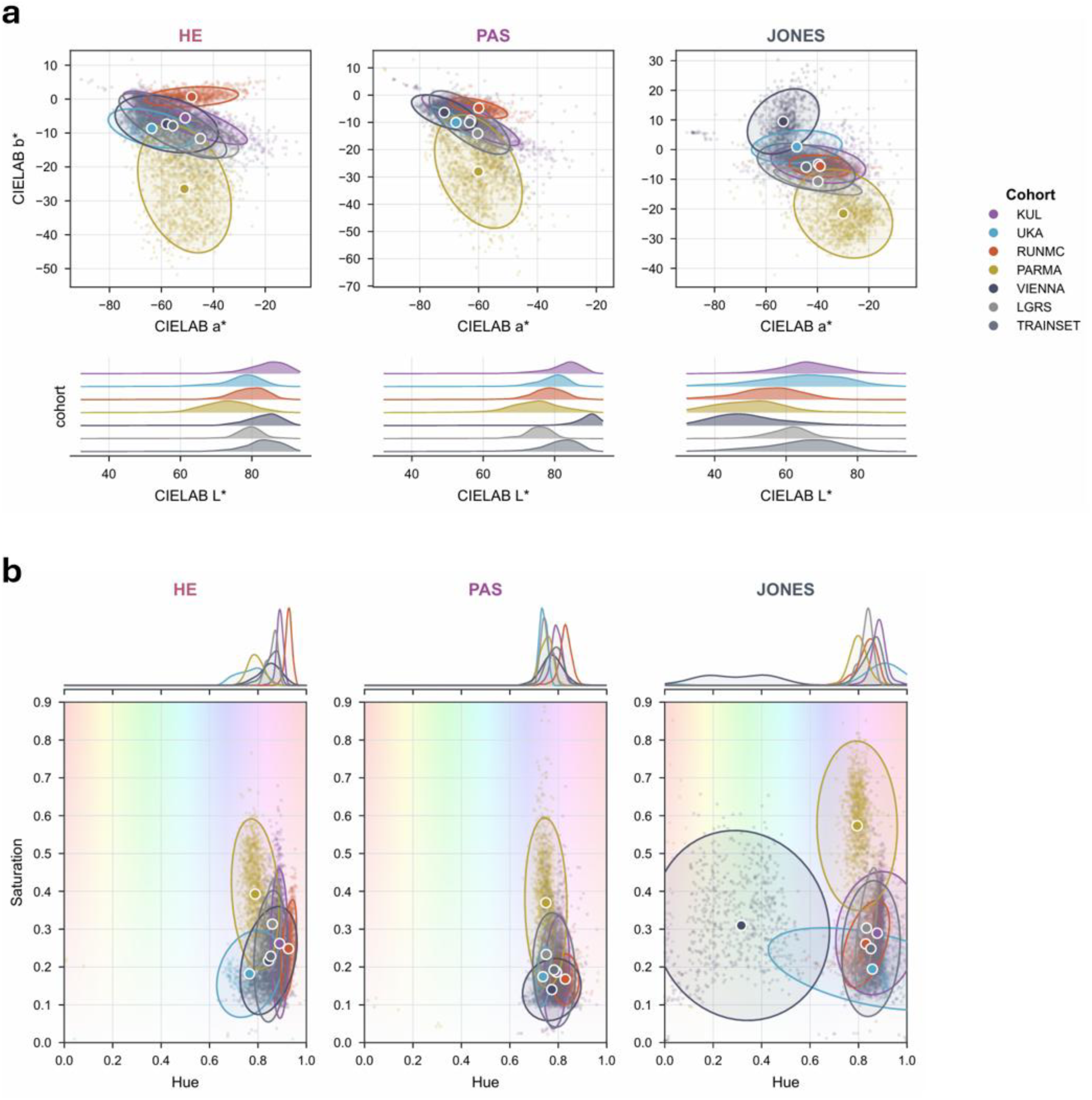
Analysis of stain variation across the train and validation cohorts. **a.** Upper panel = CIELAB chromaticity where each point is a sampled patch, the ellipse the 2SD spread and the marker the centroid of the train cohort and all the validation cohorts. **b.** Hue vs saturation for the datasets across the 3 stainings. The data indicate substantial variation between the training cohort and the validation cohorts across the 3 stainings: HE, JONES (Silver) and PAS.

**Extended Data Figure 4.**
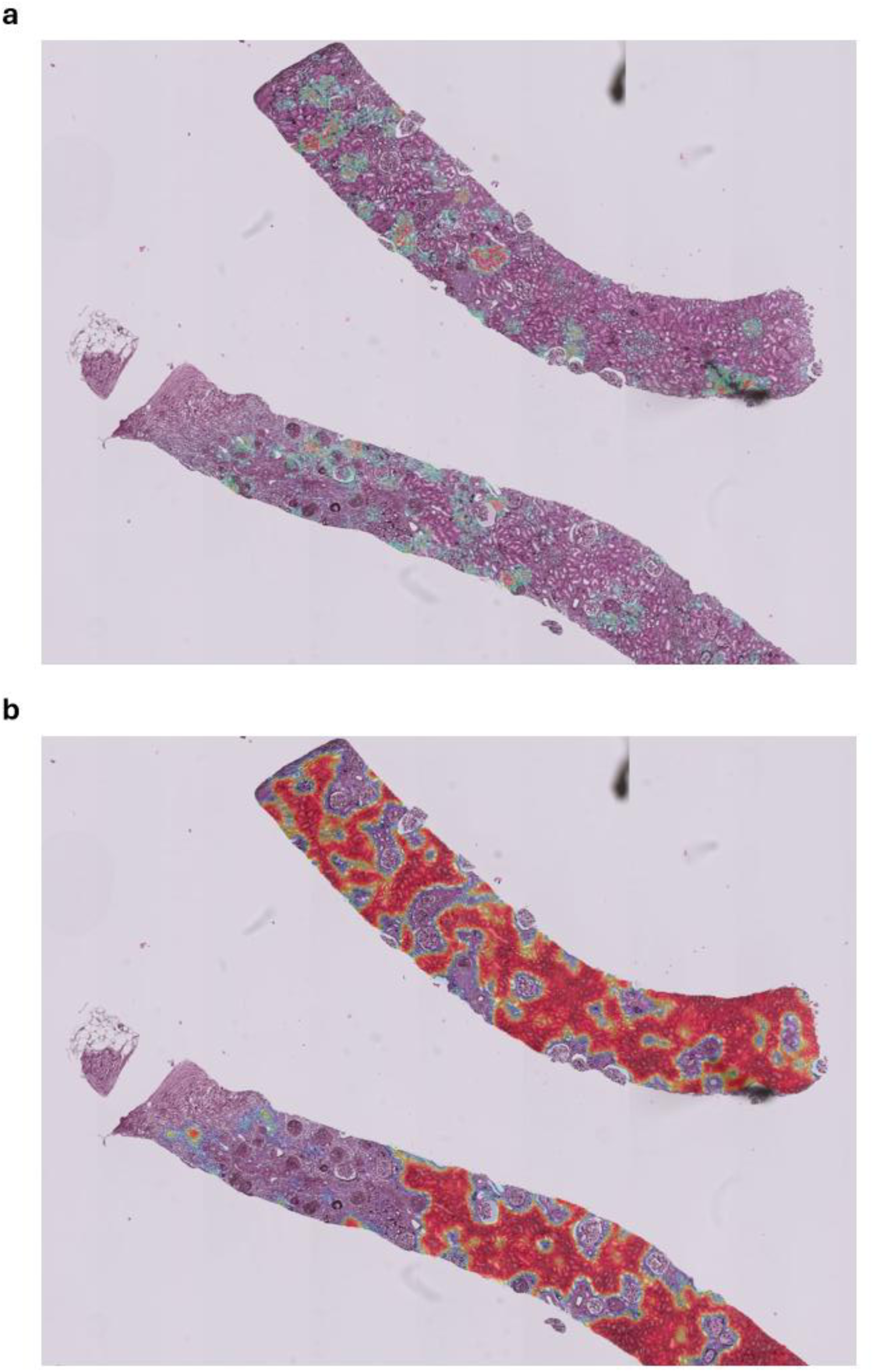
Explainability maps for peritubular capillaritis. **a.** BanffNET pixel-based explainability map for ptc showing fine-grained localization of high-probability regions. **b.** Smoothened patch-level attention heatmap for ptc of the same biopsy derived from the attention-MIL model.

**Extended Data Figure 5.**
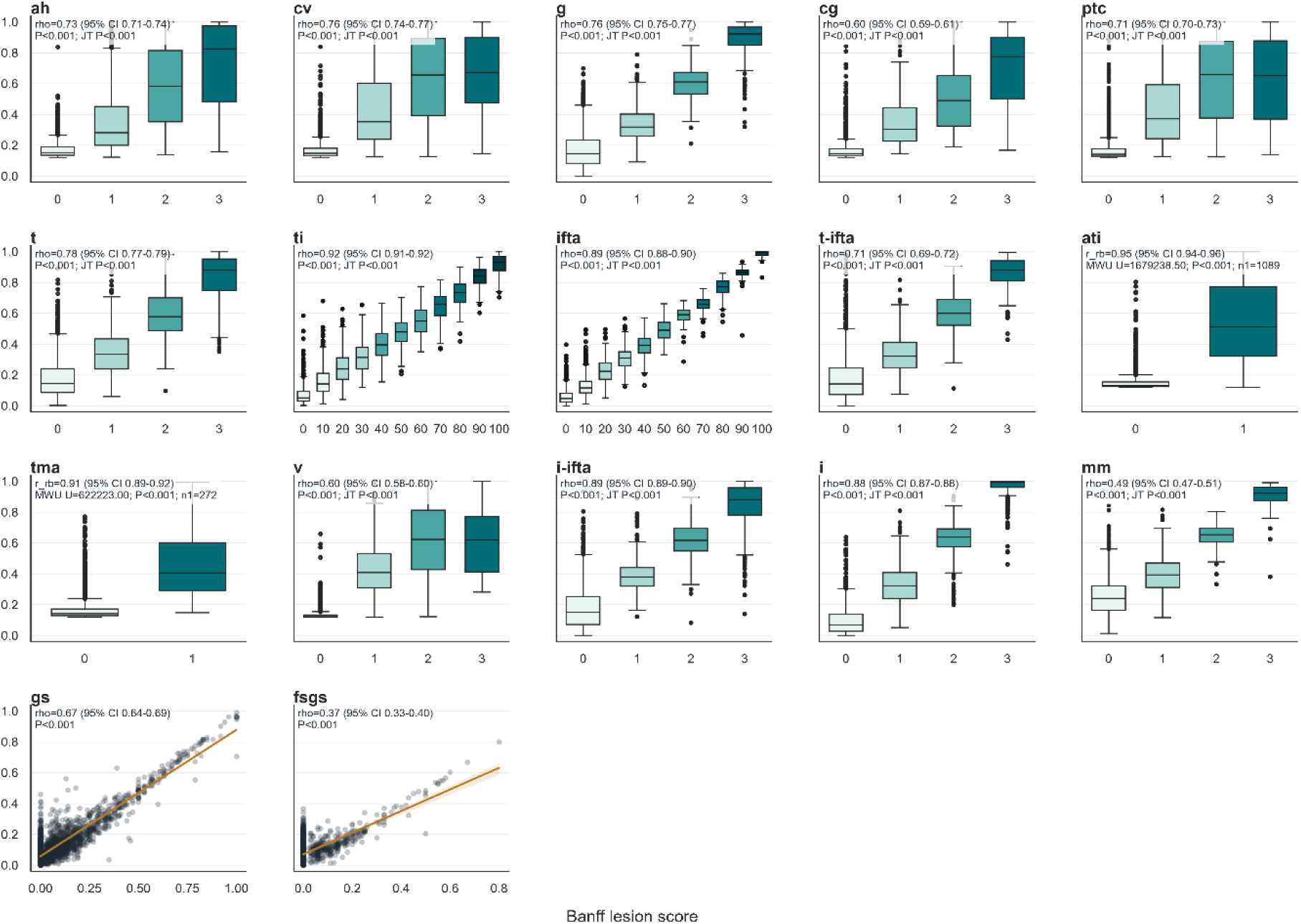
Correlations between pathologist Banff scores and BanffNET scores on the train cohort. BanffNET scores are presented as boxplots stratified by incremental ordinal pathologist Banff score. Pathologist scores for global glomerulosclerosis (gs) and focal and segmental glomerulosclerosis (fsgs) are presented as the fraction of glomeruli affected by the respective lesion as a continuous value. Binary pathologist scores are represented as rank biserial (rb) effect and tested with the non-parametric Mann-Whtiney U (MWU) test; ordinal pathologist scores are represented as Spearman rho (rho) and tested with the Jonckheere-Terpstra (JT) ordered trend test; continuous variables are tested with the Spearman test and effect size is the Spearman rho. Noteworthy, for all parameters, especially when the pathologist scored “0”, BanffNET potentially identified cases that were mislabeled, indicating potentially an increase in sensitivity for early lesion detection.

**Extended Data Figure 6.**
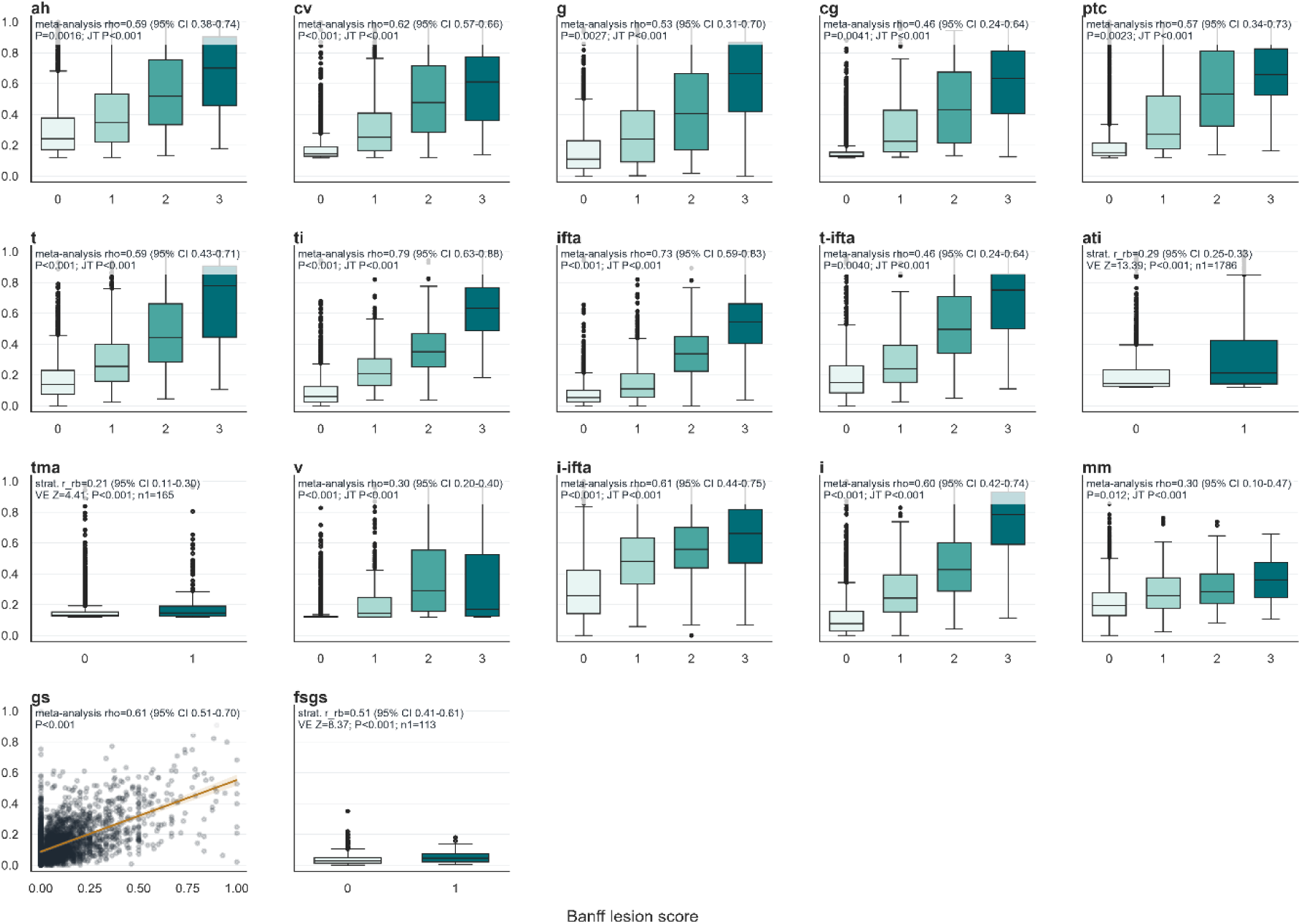
Correlations between pathologist Banff scores and BanffNET scores on the validation cohort. BanffNET scores (y-axes) are represented as boxplots or scatterplot conditioned on the incremental pathologist lesion scores (binary, ordinal or continuous, depending on the lesion). Pathologist scores for global glomerulosclerosis (gs) are presented as the fraction of glomeruli with global glomerulosclerosis as a continuous value and pathologist focal and segmental glomerulosclerosis (fsgs) scores are presented as binary variable (0 = absent, 1 = present). Coefficients and statistical tests represent meta-analysis/stratification of cohort-specific metrics with estimated of the confidence intervals with bootstrapping/permutation tests: binary pathologist scores are represented as rank biserial (rb) effect and tested with the stratified non-parametric Van Elteren (VE) test; ordinal pathologist scores are represented as Spearman rho (rho) after meta-analysis and tested with the Jonckheere-Terpstra (JT) ordered trend test; continuous variables are represented as Spearman rho (rho) after meta-analysis and tested with the Hartung-Knapp inference to estimate the standard error, confidence intervals and p-value.

**Extended Data Figure 7.**
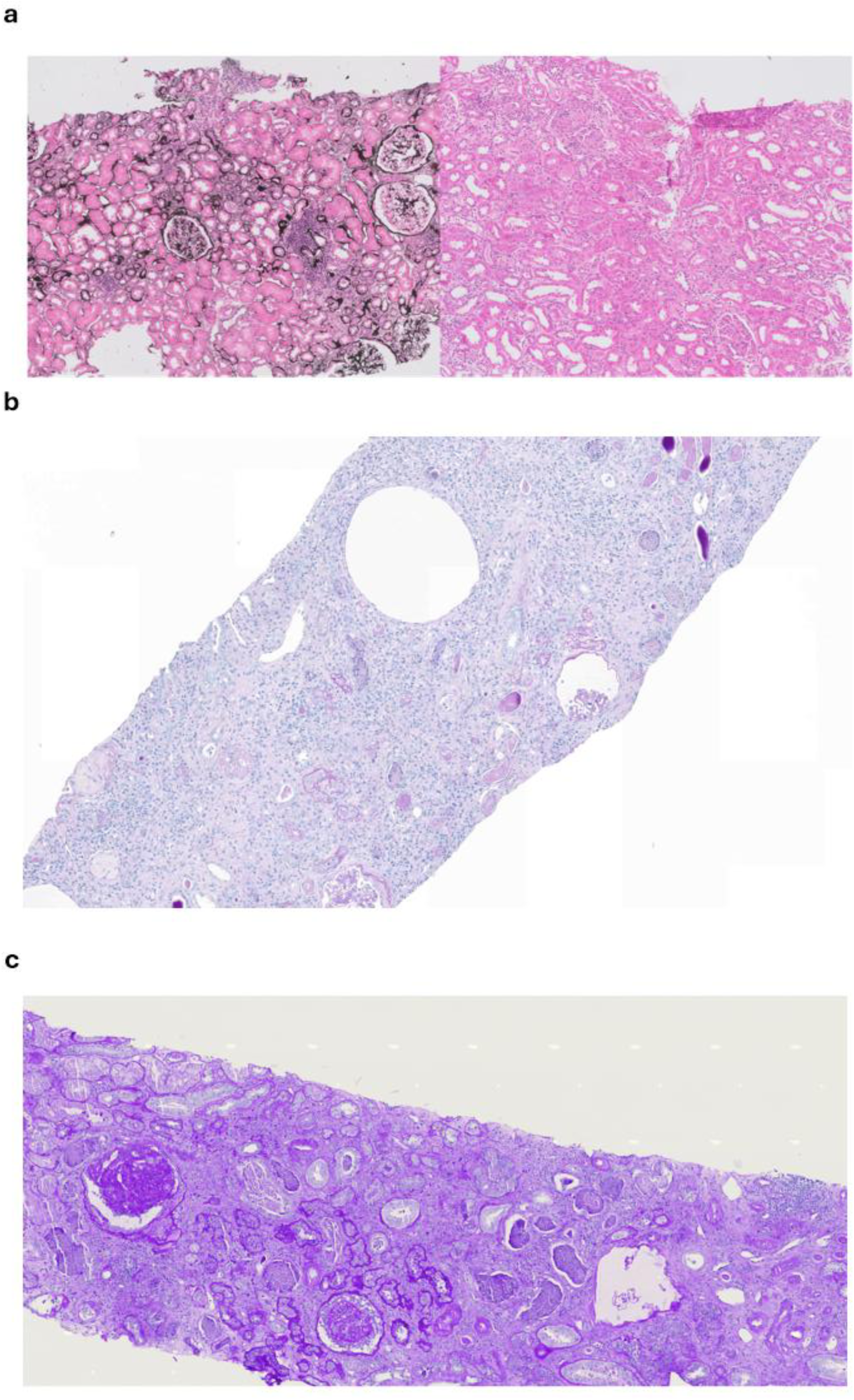
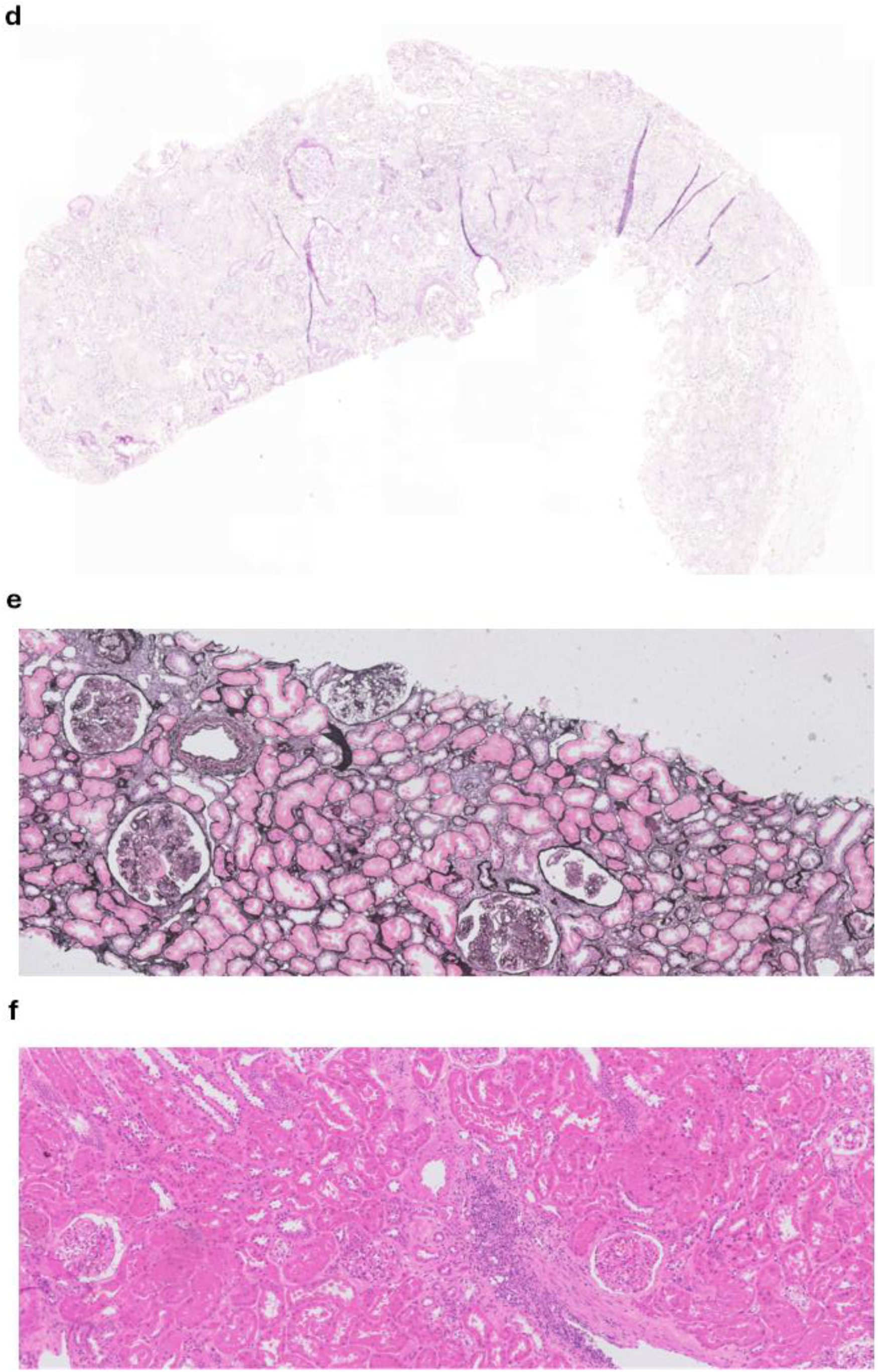
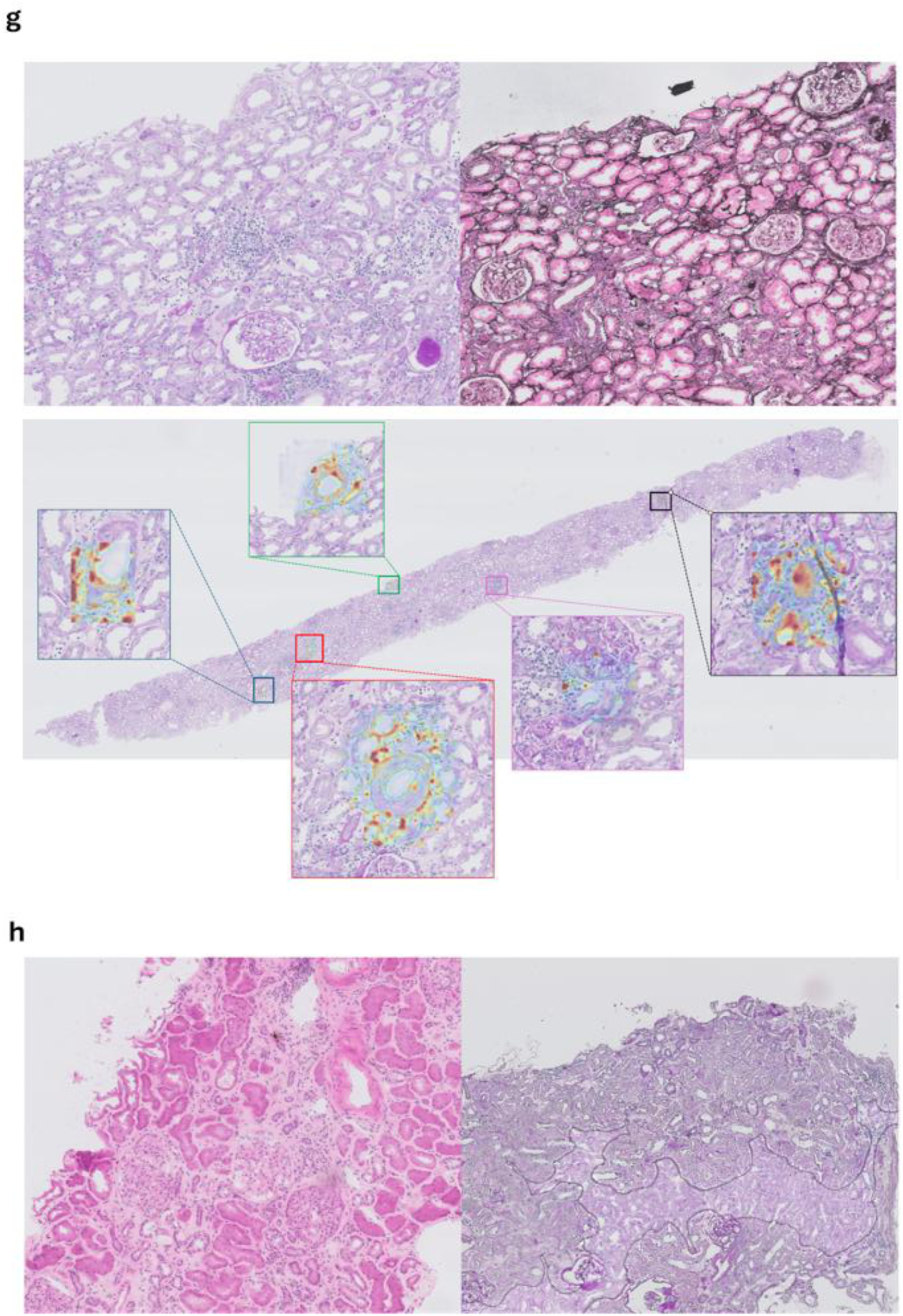
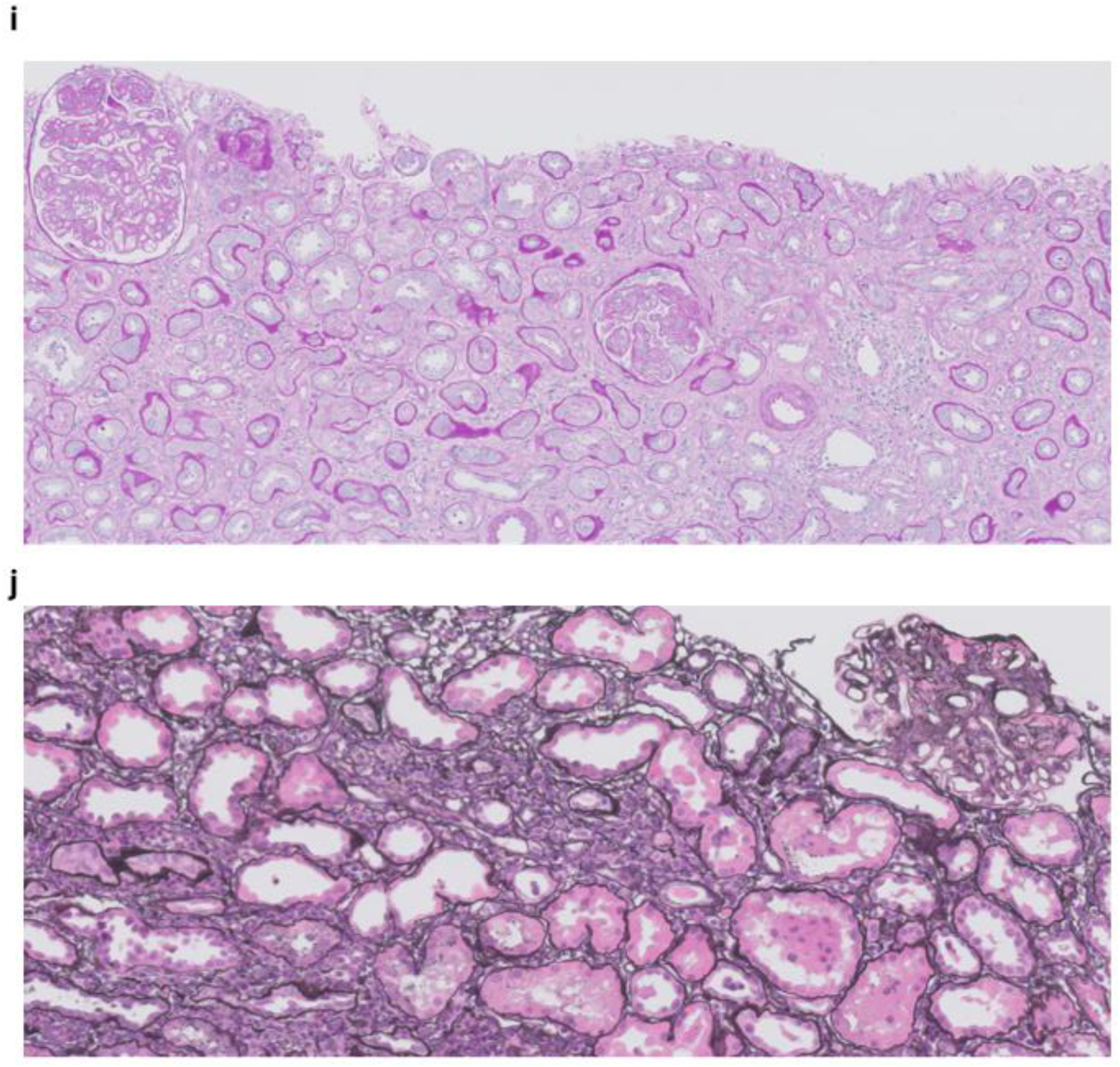
Visualizations of the validation cases with the highest overall discrepancy between the local pathologist lesion scores and the BanffNET lesion scores. **a.** Case 1: the silver staining (left) and the HCE staining (right) have a discrepant presentation. **b.** Case 2: extensive fibrosis and inflammation due to chronic active pyelonephritis. **c.** Case 3: extensive fibrosis and inflammation due to chronic-active pyelonephritis. **d.** Case 4: despite the poor quality PAS staining were the central review and the MMDx analysis more in line with BanffNET (e.g. interstitial fibrosis, tubular atrophy and double contours). **e.** Case 5: recurrence of proliferative glomerulonephritis, central reviews are more in line with BanffNET than with the original local pathologist score (e.g. presence of glomerulitis, tma and mesangial matrix increase). **f.** Case 6: HCE staining mislabeled as PAS. BanffNET correctly applied to the HCE staining indicated high v score (0.94), in line with central review where 2/3 pathologists score v2, whereas the local pathologist scored v0. **g.** Case 7: the PAS and silver staining were mislabeled. Corrected reanalysis of BanffNET indicated e.g. high ati score in both PAS and silver, in line with local review and one of the central reviews. BanffNET on the PAS staining was high for endothelitis (v) whereas all pathologists scored v0. Interestingly, attention visualization showed intense heatmaps in and around vessels exclusively. The patient lost the graft <3 months after the biopsy. **h.** Case 8: the HCE staining was done on a frozen section whereas the PAS staining showed many artifacts (trapped air bubbles under the cover slip, out-of-focus), influencing the assessment by BanffNET. Indeed, BanffNET on the silver staining was more in line with the local and central pathologist reviews. **i.** Case 9: recurrence of proliferative glomerulonephritis with glomerular scores more in line with the central review. **j.** Case 10: scores are more in line with central reviews than local review (e.g. interstitial fibrosis and tubular atrophy scores). For in-depth description of case discrepancies between BanffNET scores, original local pathologist review and blinded central pathologist reviews (3 pathologists), see **Extended Data Tables 8-17**.

**Extended Data Figure 8.**
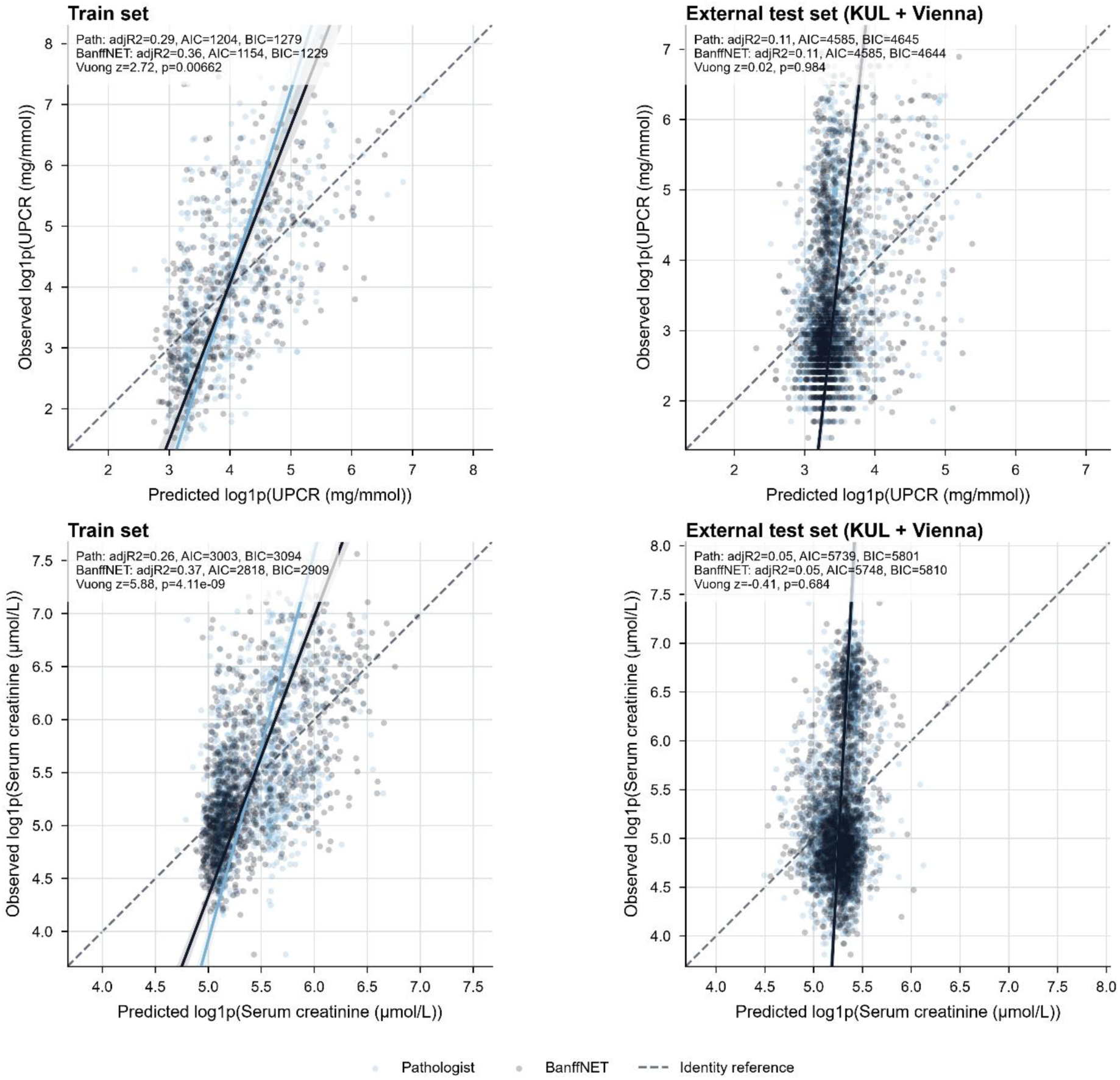
Comparison of multivariable linear regression models consisting of individual BanffNET or pathologist Banff lesion scores for urine protein-creatinine ratio (UPCR) or serum creatinine (SCr) at time of biopsy. Multivariable models consisting of either BanffNET lesion scores or pathologist Banff lesion scores and predicting the log UPCR (upper panels) and the log SCr (lower panels) show significantly higher explained variation by BanffNET lesion scores when fitted to the train cohort (adjusted R^2^ 0.36 vs. 0.29, resp. for UPCR and adjusted R^2^ 0.37 vs 0.26, resp. for SCr) with a higher closeness to the true data in favor of the multivariable BanffNET model (Vuong closeness test for comparison of non-nested models, Z = +2.72, P = 0.007 for UPCR and Z = +5.88, P <0.0001 for SCr), whereas no significant difference was observed when fitted on the external validation set (right panels).

**Extended Data Figure 9.**
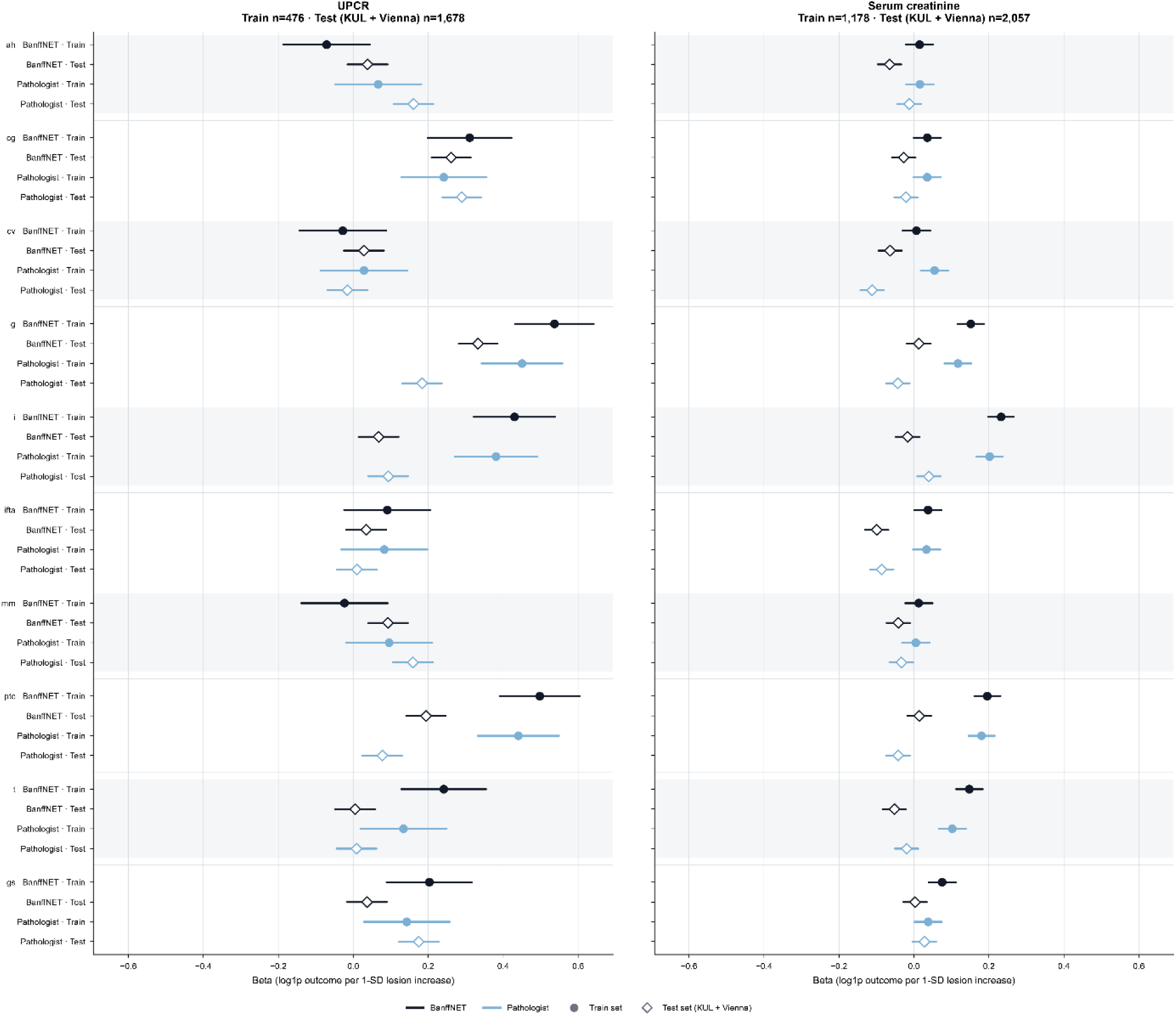
Comparison of individual BanffNET and pathologist Banff lesion scores associations with death-censored graft failure in the training and validation cohorts. Individual BanffNET and pathologist Banff lesion scores separately fitted on the training cohort and the validation cohort for urine protein-creatinine ratio (left panel) and serum creatinine (right panel). The association between the individual lesion scores and both outcome measures, whether this is BanffNET scores or pathologist Banff scores, is mostly determined by cohort characteristics. Data are represented as z-score normalized per SD increase in the scores for the log-transformed outcome measure.

**Extended Data Figure 10.**
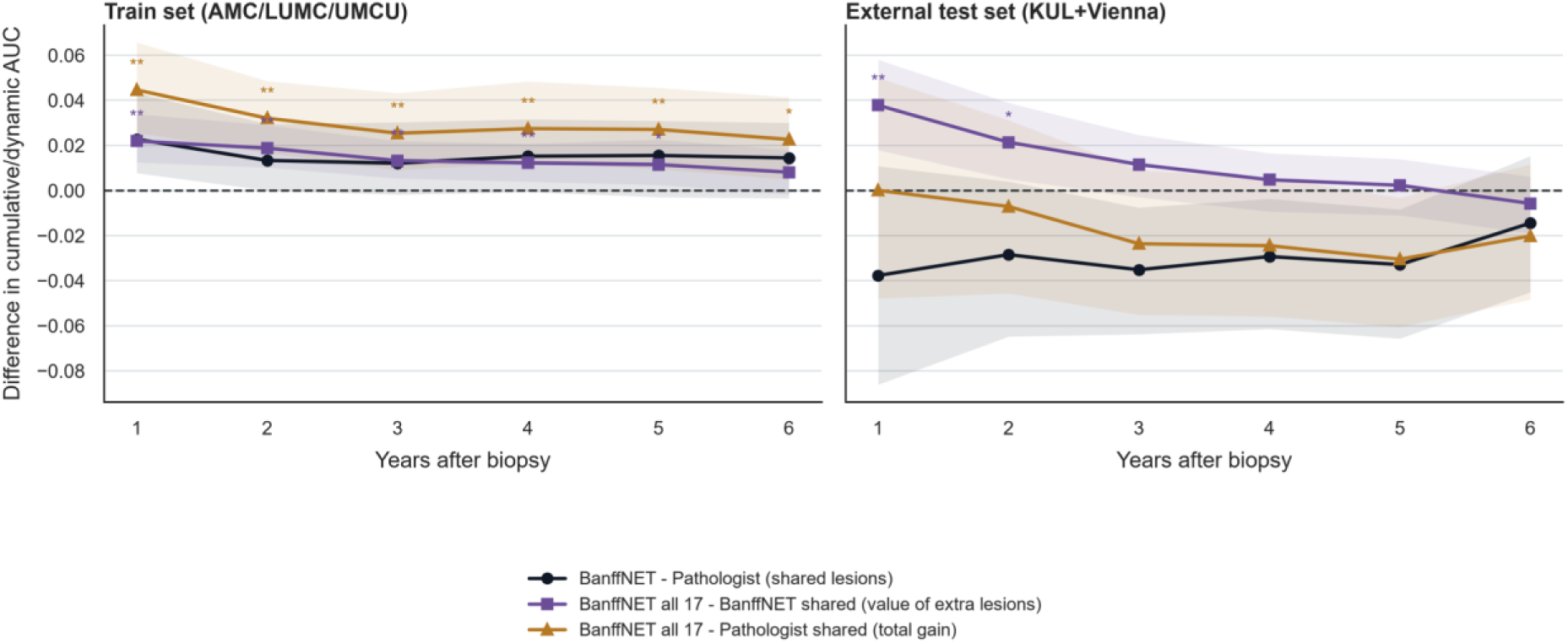
Differences in cumulative-dynamic ROC curve analysis for the discrimination of death-censored graft failure in the time after biopsy. Differences in cumulative-dynamic ROC curve analysis in the train cohort (left panel) and the validation cohort (right panel). Positive values favor the first-named model. Paired permutation test on within-patient score swaps; BH-adjusted across horizons. ***q<0.001, **q<0.01, *q<0.05; unmarked horizons are not significant.

**Extended Data Figure 11.**
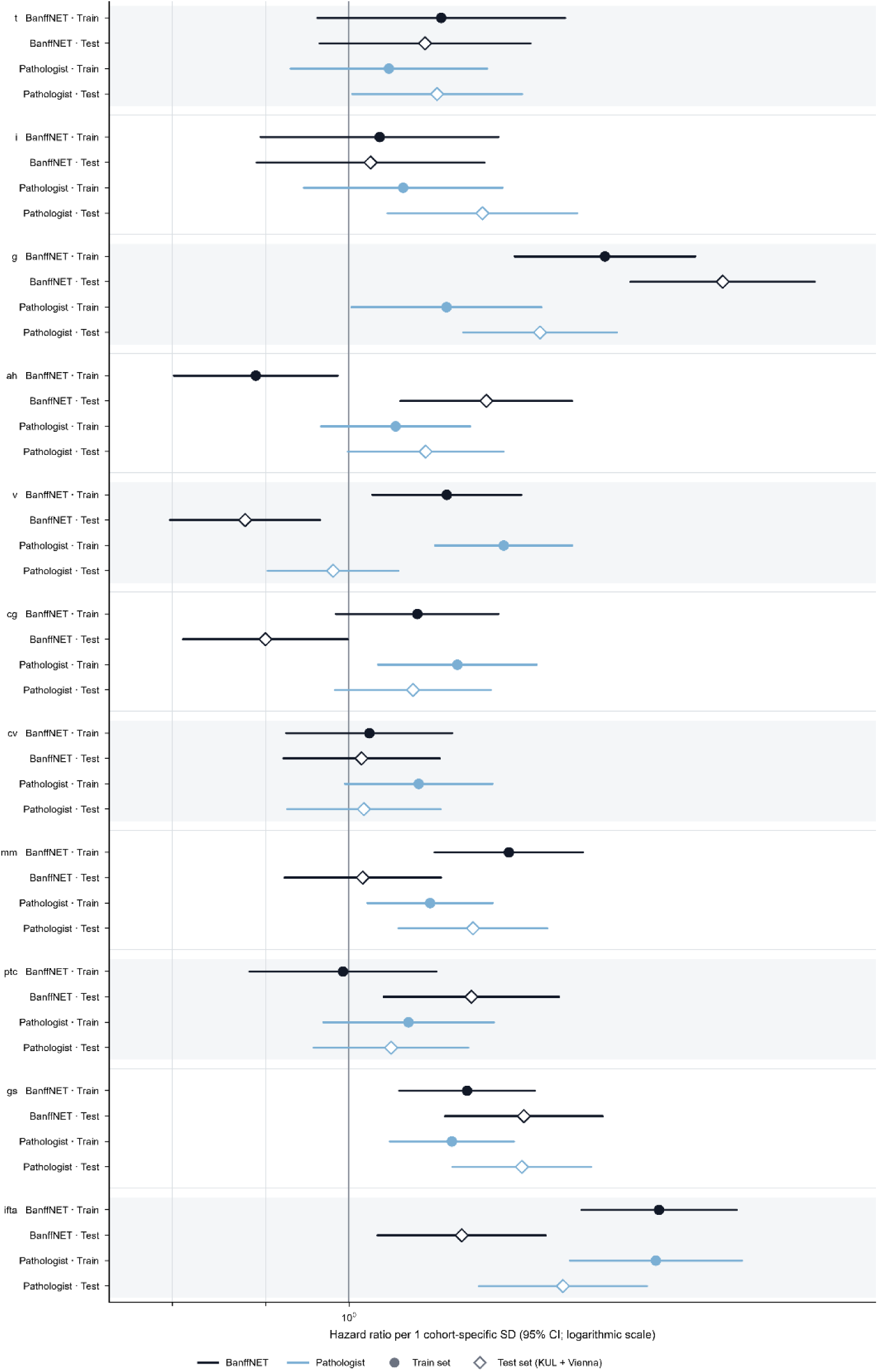
Comparison of individual BanffNET and pathologist Banff lesion score associations with death-censored graft failure in the training and validation cohorts. Individual BanffNET and pathologist Banff lesion scores separately fitted on the training cohort and the validation cohort for the association with death-censored graft failure (DCGF). Data are represented as hazard ratio per z-score normalized SD increase in the scores for time to DCGF.

**Extended Data Figure 12.**
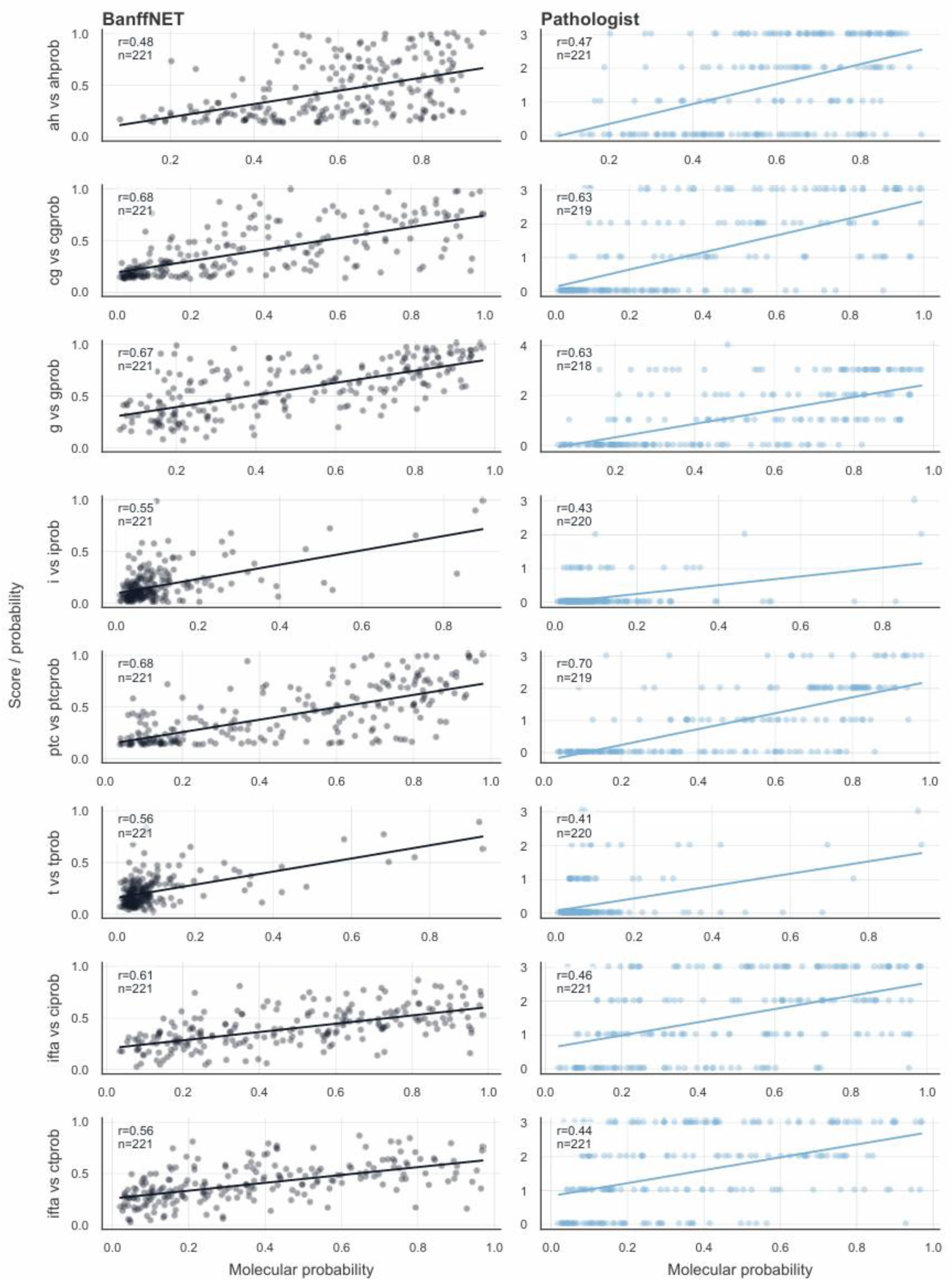
Correlations of pathologist Banff lesion scores and BanffNET lesion scores with histological lesion probabilities defined by MMDx. Individual biopsy dotplots with linear regression lines indicating an overall stronger correlation between BanffNET lesion scores and corresponding MMDx lesion probabilities than pathologist Banff lesion scores and corresponding MMDx lesion probabilities. Only the pathologist ptc score showed a marginally better model fit.

**Extended Data Table 1.**
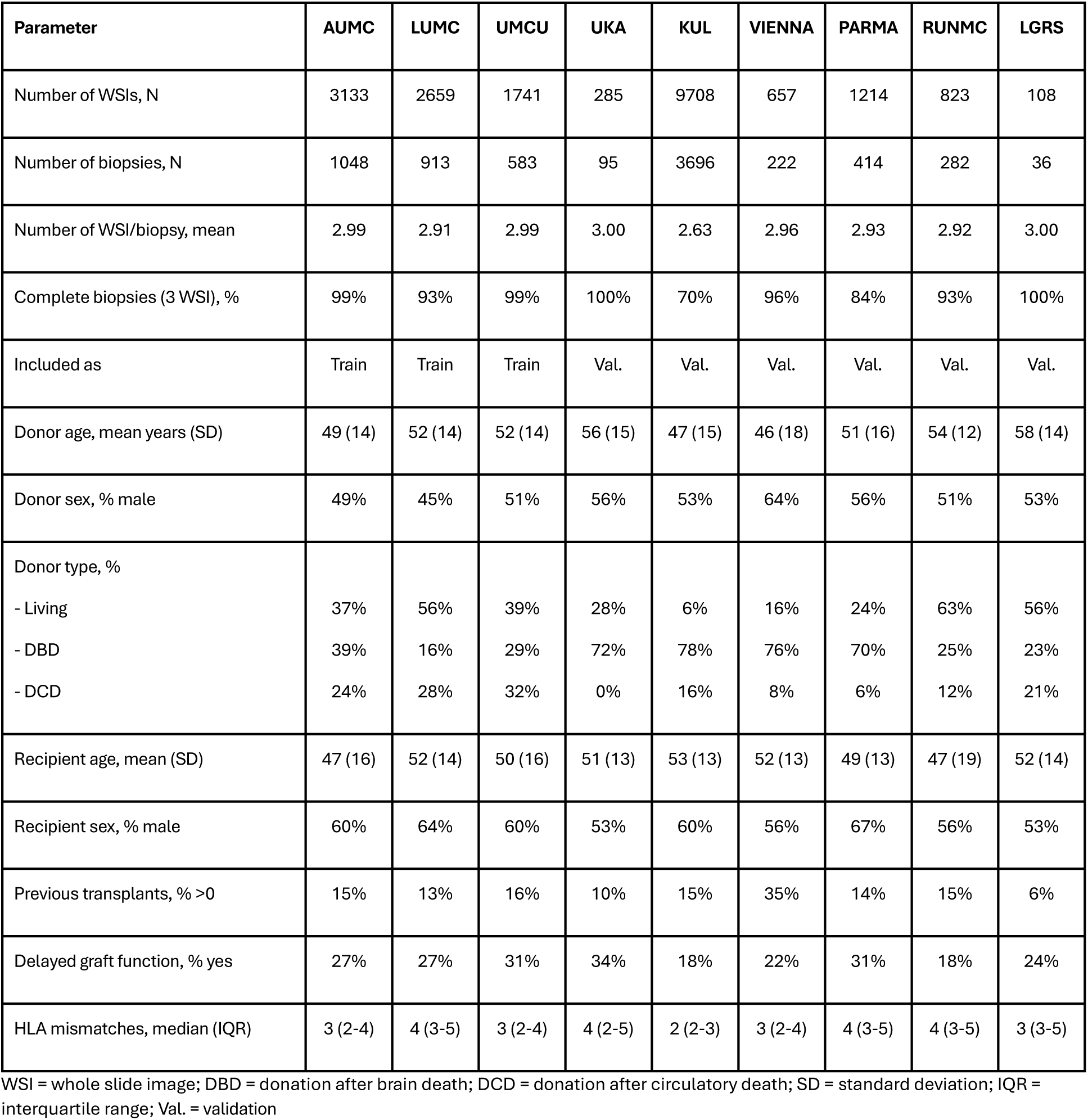
Baseline characteristics of the included cohorts.

| Parameter | AUMC | LUMC | UMCU | UKA | KUL | VIENNA | PARMA | RUNMC | LGRS |
| --- | --- | --- | --- | --- | --- | --- | --- | --- | --- |
| Number of WSIs, N | 3133 | 2659 | 1741 | 285 | 9708 | 657 | 1214 | 823 | 108 |
| Number of biopsies, N | 1048 | 913 | 583 | 95 | 3696 | 222 | 414 | 282 | 36 |
| Number of WSI/biopsy, mean | 2.99 | 2.91 | 2.99 | 3.00 | 2.63 | 2.96 | 2.93 | 2.92 | 3.00 |
| Complete biopsies (3 WSI), % | 99% | 93% | 99% | 100% | 70% | 96% | 84% | 93% | 100% |
| Included as | Train | Train | Train | Val. | Val. | Val. | Val. | Val. | Val. |
| Donor age, mean years (SD) | 49 (14) | 52 (14) | 52 (14) | 56 (15) | 47 (15) | 46 (18) | 51 (16) | 54 (12) | 58 (14) |
| Donor sex, % male | 49% | 45% | 51% | 56% | 53% | 64% | 56% | 51% | 53% |
| Donor type, % |  |  |  |  |  |  |  |  |  |
| - Living | 37% | 56% | 39% | 28% | 6% | 16% | 24% | 63% | 56% |
| - DBD | 39% | 16% | 29% | 72% | 78% | 76% | 70% | 25% | 23% |
| - DCD | 24% | 28% | 32% | 0% | 16% | 8% | 6% | 12% | 21% |
| Recipient age, mean (SD) | 47 (16) | 52 (14) | 50 (16) | 51 (13) | 53 (13) | 52 (13) | 49 (13) | 47 (19) | 52 (14) |
| Recipient sex, % male | 60% | 64% | 60% | 53% | 60% | 56% | 67% | 56% | 53% |
| Previous transplants, % >0 | 15% | 13% | 16% | 10% | 15% | 35% | 14% | 15% | 6% |
| Delayed graft function, % yes | 27% | 27% | 31% | 34% | 18% | 22% | 31% | 18% | 24% |
| HLA mismatches, median (IQR) | 3 (2-4) | 4 (3-5) | 3 (2-4) | 4 (2-5) | 2 (2-3) | 3 (2-4) | 4 (3-5) | 4 (3-5) | 3 (3-5) |

WSI = whole slide image; DBD = donation after brain death; DCD = donation after circulatory death; SD = standard deviation; IQR = interquartile range; Val. = validation

**Extended Data Table 2.** Overview and distribution of Banff lesion scores for the included cohorts.

| Score | AUMC | LUMC | UMCU | UKA | KUL | VIENNA | PARMA | RUNMC | LGRS |
| --- | --- | --- | --- | --- | --- | --- | --- | --- | --- |
| tma | 6% | 16% | 8% | 12% | 3% | n/a | 5% | 14% | 0% |
| ati | 23% | 55% | 43% | 49% | 36% | n/a | 6% | 74% | 64% |
| g | 0 (0-1) | 0 (0-2) | 0 (0-1) | 0 (0-0) | 0 (0-0) | 1 (0-2) | 0 (0-0) | 0 (0-1) | 0 (0-1) |
| cg | 0 (0-0) | 0 (0-0) | 0 (0-0) | 0 (0-0) | 0 (0-0) | 0 (0-2) | 0 (0-0) | 0 (0-0) | 0 (0-0) |
| mm | 0 (0-0) | 0 (0-0) | 0 (0-0) | 0 (0-0) | 0 (0-0) | 0 (0-1) | 0 (0-0) | 0 (0-0) | 0 (0-0) |
| i | 0 (0-1) | 1 (0-2) | 1 (0-1) | 1 (0-2) | 0 (0-0) | 0 (0-0) | 0 (0-0) | 0 (0-2) | 0 (0-1) |
| t | 0 (0-1) | 1 (0-2) | 1 (0-1) | 0 (0-1) | 0 (0-1) | 0 (0-0) | 0 (0-1) | 1 (0-1) | 0 (0-1) |
| ti<br>(ord.) | 1 (1-3) | 2 (1-2) | 1 (1-2) | 1 (1-3) | n/a | 1 (0-1) | 0 (0-1) | 1 (0-2) | 1 (1-2) |
| ti<br>(cont.) | 20 (10-60) | 30 (10-50) | 20 (10-40) | 20 (10-60) | n/a | n/a | n/a | 20 (0-50) | 20 (10-40) |
| ifta<br>(ord.) | 1 (0-2) | 1 (0-1) | 1 (0-2) | 1 (0-1) | 1 (1-1) | 2 (0-3) | 1 (0-1) | 1 (0-2) | 1 (1-2) |
| ifta<br>(cont.) | 10 (0-40) | 10 (0-20) | 10 (0-30) | 10 (0-20) | n/a | n/a | n/a | 10 (0-30) | 10 (10-30) |
| i-ifta | 0 (0-3) | 1 (0-3) | 1 (0-2) | 1 (0-2) | n/a | 0 (01) | 1 (0-1) | 1 (0-3) | 1 (0-2) |
| t-ifta | 0 (0-1) | 0 (0-1) | 1 (0-1) | 0 (0-1) | n/a | 0 (0-0) | 0 (0-0) | 0 (0-1) | 0 (0-1) |
| ptc | 0 (0-0) | 0 (0-1) | 0 (0-1) | 0 (0-1) | 0 (0-0) | 0 (0-2) | 0 (0-0) | 1 (0-1) | 0 (0-1) |
| v | 0 (0-0) | 0 (0-0) | 0 (0-0) | 0 (0-0) | 0 (0-0) | 0 (0-0) | 0 (0-0) | 0 (0-0) | 0 (0-0) |
| cv | 0 (0-1) | 1 (0-1) | 1 (0-1) | 1 (0-2) | 1 (0-1) | 1 (0-2) | 0 (0-1) | 1 (0-2) | 1 (1-2) |
| ah | 0 (0-1) | 1 (0-1) | 1 (0-1) | 1 (0-1) | 0 (0-1) | 2 (0-3) | 0 (0-1) | 1 (1-2) | 1 (0-2) |

ord. = scored in an ordinal way (according to the latest Banff classification); cont. = scored in a continuous, more granular way on a scale from 0-100% with incremental steps of 10%; n/a = not available. Data are presented as median (interquartile range).

**Extended Data Table 3.** Mapping of the histological lesion scores to probability space to train BanffNET.

| Score | Type | Scale | Probability mapping |
| --- | --- | --- | --- |
| tma | Binary | absent/present | 0.0; 1.0 |
| ati | Binary | absent/present | 0.0; 1.0 |
| g | Ordinal | 0-3 | 0.0; 0.33; 0.67; 1.0 |
| cg<sup>1</sup> | Ordinal | 0-3 | 0.0; 0.33; 0.67; 1.0 |
| mm | Ordinal | 0-3 | 0.0; 0.33; 0.67; 1.0 |
| i | Ordinal | 0-3 | 0.0; 0.33; 0.67; 1.0 |
| t | Ordinal | 0-3 | 0.0; 0.33; 0.67; 1.0 |
| ti | Ordinal | 0-100% (+10%) | 0.0; 0.1; 0.2; 0.3; 0.4; 0.5; 0.6; 0.7; 0.8; 0.9; 1.0 |
| ifta | Ordinal | 0-100% (+10%) | 0.0; 0.1; 0.2; 0.3; 0.4; 0.5; 0.6; 0.7; 0.8; 0.9; 1.0 |
| i-ifta | Ordinal | 0-3 | 0.0; 0.33; 0.67; 1.0 |
| t-ifta | Ordinal | 0-3 | 0.0; 0.33; 0.67; 1.0 |
| ptc | Ordinal | 0-3 | 0.0; 0.33; 0.67; 1.0 |
| v | Ordinal | 0-3 | 0.0; 0.33; 0.67; 1.0 |
| cv | Ordinal | 0-3 | 0.0; 0.33; 0.67; 1.0 |
| ah | Ordinal | 0-3 | 0.0; 0.33; 0.67; 1.0 |
| gs | Continuous | 0-100% | 0.0 – 1.0 |
| fsgs | Continuous | 0-100% | 0.0 – 1.0 |

NIP = Noisy-OR pooling; GIP = Gated Instance Pooling

**Extended Data Table 4.** Model architecture and parameter count comparison between state-of-the-art multiple instance learning models and NIP/GIP aggregation models.

| Model | Number of parameters | Configuration |
| --- | --- | --- |
| ABMIL<sup>32</sup> | 788226 | Shallow attention-based MIL with gated attention, small network size<br>Input projection: 1024 → 512 (ReLU)<br>Attention module: Gated (Tanh ⊗ Sigmoid → Linear → 1)<br>Aggregation: Softmax-weighted average of patch embeddings<br>Classifier: 512 → 1 (Sigmoid) |
| TransMIL<sup>18</sup> | 2671633 | Transformer-based MIL with Nystrom self-attention.<br>Input projection: 1024 → 512 (ReLU)<br>Transformer encoder: 2 × NystromAttention blocks (8 heads, 512 dim)<br>Positional encoding: PPEG (depth-wise convolutions: 7×7, 5×5, 3×3)<br>Aggregation: CLS token<br>Classifier: 512 → 1 (Sigmoid) |
| BanffNET-NIP | 1025 | No hyperparameters tuning required |
| BanffNET-GIP | 2050 | No hyperparameters tuning required |

**Extended Data Table 5.** Comparison of performance between state-of-the-art attention-based multiple instance learning models and NIP/GIP aggregation models.

| Model | ati | ifta | ptc | t | Mean |
| --- | --- | --- | --- | --- | --- |
| ABMIL | 0.49 | 0.84 | 0.85 | 0.80 | **0.75** |
| TransMIL | 0.4 | 0.91 | 0.68 | 0.81 | **0.70** |
| BanffNET-NIP/GIP | 0.53 | 0.91 | 0.91 | 0.92 | **0.82** |

Data are represented as Pearson correlation coefficient with the Leiden Global Reader Study consensus

**Extended Data Table 6.** Correlations between BanffNET lesion scores and the Pathologist Banff lesion scores.

| Lesion | Training data | validation data |
| --- | --- | --- |
| tma | rb = 0.91 (0.89-0.92), P<0.001 | rb<sub>strat</sub> = 0.21 (0.11-0.30), P<0.001 |
| ati | rb = 0.95 (0.94-0.96), P<0.001 | rb<sub>strat</sub> = 0.29 (0.25-0.33), P<0.001 |
| g | rho = 0.76 (0.75-0.77), P<0.001 | rho<sub>meta</sub> = 0.53 (0.31-0.70), P<0.001 |
| cg | rho = 0.60 (0.59-0.61), P<0.001 | rho<sub>meta</sub> = 0.46 (0.24-0.64), P<0.001 |
| mm | rho = 0.49 (0.47-0.51), P<0.001 | rho<sub>meta</sub> = 0.30 (0.10-0.47), P<0.001 |
| i | rho = 0.88 (0.87-0.88), P<0.001 | rho<sub>meta</sub> = 0.60 (0.42-0.74), P<0.001 |
| t | rho = 0.78 (0.77-0.79), P<0.001 | rho<sub>meta</sub> = 0.59 (0.43-0.71), P<0.001 |
| ti | rho = 0.92 (0.91-0.92), P<0.001 | rho<sub>meta</sub> = 0.79 (0.63-0.88), P<0.001 |
| ifta | rho = 0.89 (0.88-0.90), P<0.001 | rho<sub>meta</sub> = 0.73 (0.59-0.83), P<0.001 |
| i-ifta | rho = 0.89 (0.89-0.90), P<0.001 | rho<sub>meta</sub> = 0.61 (0.44-0.75), P<0.001 |
| t-ifta | rho = 0.71 (0.69-0.72), P<0.001 | rho<sub>meta</sub> = 0.46 (0.24-0.64), P<0.001 |
| ptc | rho = 0.71 (0.70-0.73), P<0.001 | rho<sub>meta</sub> = 0.57 (0.34-0.73), P<0.001 |
| v | rho = 0.60 (0.58-0.60), P<0.001 | rho<sub>meta</sub> = 0.30 (0.20-0.40), P<0.001 |
| cv | rho = 0.76 (0.74-0.77), P<0.001 | rho<sub>meta</sub> = 0.62 (0.57-0.66), P<0.001 |
| ah | rho = 0.73 (0.71-0.74), P<0.001 | rho<sub>meta</sub> = 0.59 (0.38-0.74), P<0.001 |
| gs | rho = 0.67 (0.64-0.69), P<0.001 | rho<sub>meta</sub> = 0.61 (0.51-0.70), P<0.001 |
| fsgs | rho = 0.37 (0.33-0.40), P<0.001 | rb<sub>strat</sub> = 0.51 (0.41-0.61), P<0.001 |

rho = Spearman rho; meta = meta-analysis; rb = rank biserial effect; strat = stratified cohort analysis. P-value for binary parameters calculated with the Mann Whitney U test (train cohort) or the Van Elteren test for cohort stratification (validation cohorts), and for ordinal parameters calculated with the Jonckheere-Terpstra ordered-trend test.

**Extended Data Table 7.**
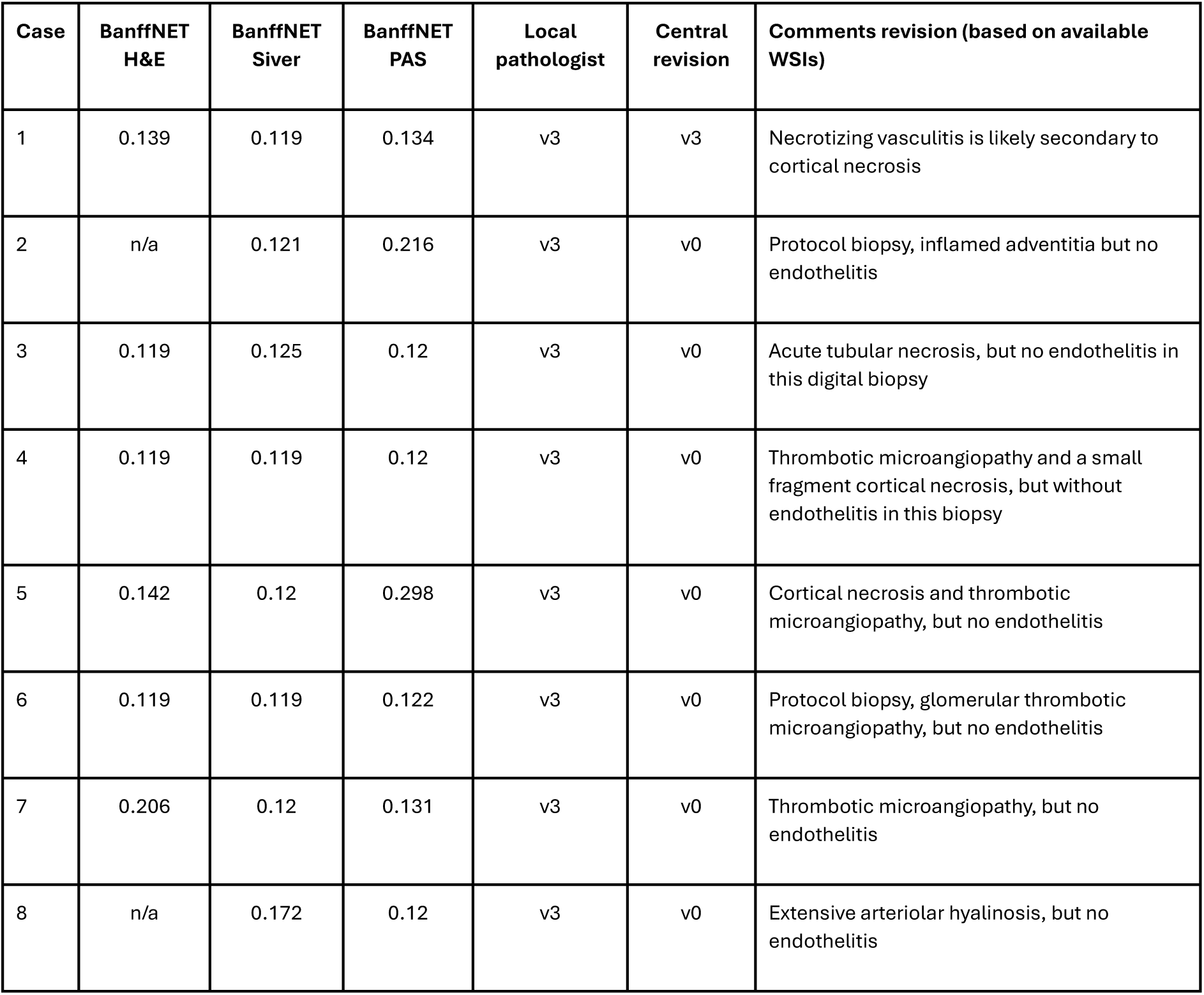
Blinded revision of validation cases with local pathologist necrotizing arteritis and low BanffNET v lesion scores.

| Case | BanffNET H&E | BanffNET Silver | BanffNET PAS | Local pathologist | Central revision | Comments revision (based on available WSIs) |
| --- | --- | --- | --- | --- | --- | --- |
| 1 | 0.139 | 0.119 | 0.134 | v3 | v3 | Necrotizing vasculitis is likely secondary to cortical necrosis |
| 2 | n/a | 0.121 | 0.216 | v3 | v0 | Protocol biopsy, inflamed adventitia but no endothelitis |
| 3 | 0.119 | 0.125 | 0.12 | v3 | v0 | Acute tubular necrosis, but no endothelitis in this digital biopsy |
| 4 | 0.119 | 0.119 | 0.12 | v3 | v0 | Thrombotic microangiopathy and a small fragment cortical necrosis, but without endothelitis in this biopsy |
| 5 | 0.142 | 0.12 | 0.298 | v3 | v0 | Cortical necrosis and thrombotic microangiopathy, but no endothelitis |
| 6 | 0.119 | 0.119 | 0.122 | v3 | v0 | Protocol biopsy, glomerular thrombotic microangiopathy, but no endothelitis |
| 7 | 0.206 | 0.12 | 0.131 | v3 | v0 | Thrombotic microangiopathy, but no endothelitis |
| 8 | n/a | 0.172 | 0.12 | v3 | v0 | Extensive arteriolar hyalinosis, but no endothelitis |

**Extended Data Table 8.**
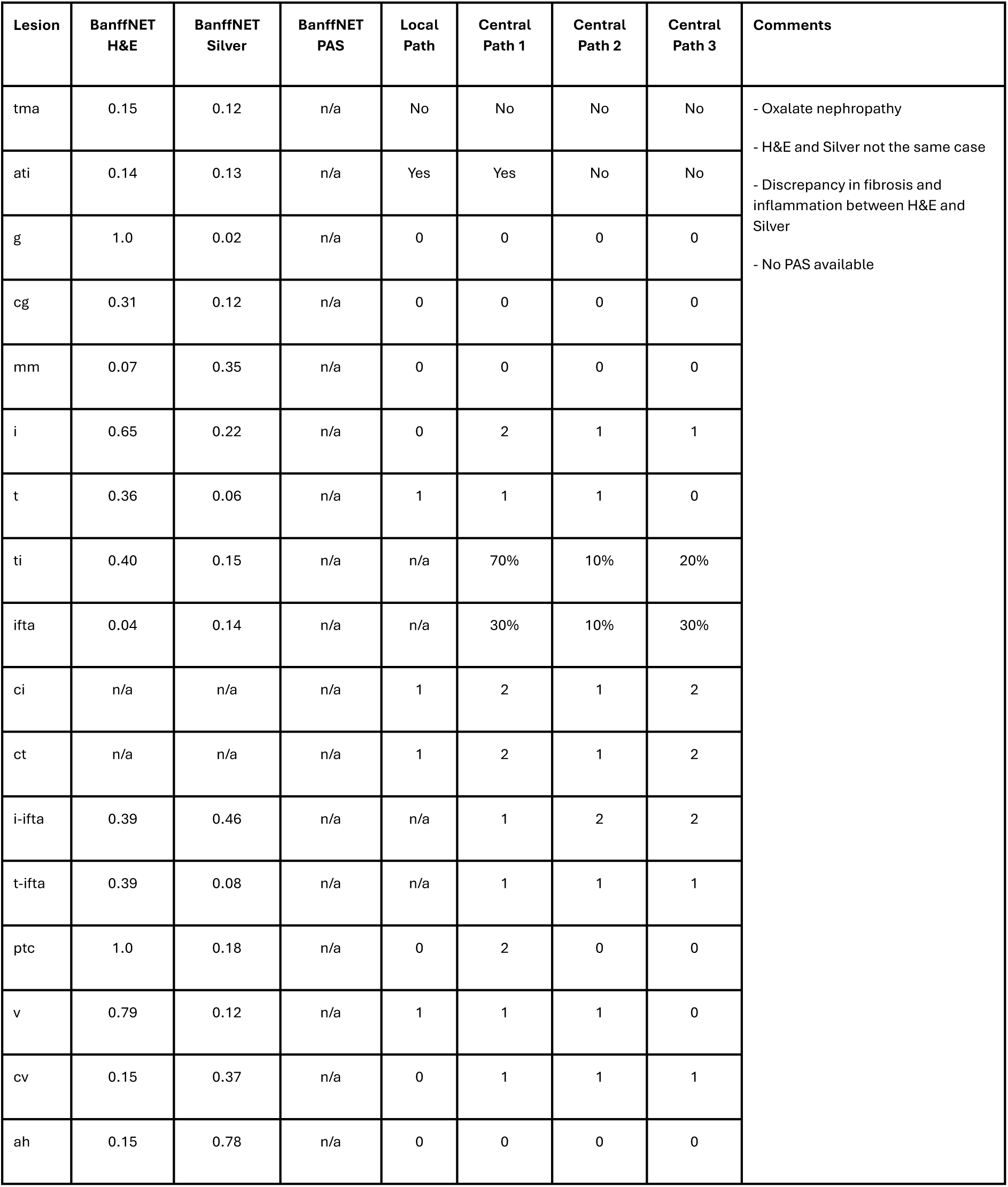
In-depth analysis of top 10 validation set cases with the highest discrepancy between local pathologist assessment and BanffNET scores across lesions (Case 1).

| Lesion | BanffNET H&E | BanffNET Silver | BanffNET PAS | Local Path | Central Path 1 | Central Path 2 | Central Path 3 | Comments |
| --- | --- | --- | --- | --- | --- | --- | --- | --- |
| tma | 0.15 | 0.12 | n/a | No | No | No | No | - Oxalate nephropathy<br>- H&E and Silver not the same case<br>- Discrepancy in fibrosis and inflammation between H&E and Silver<br>- No PAS available |
| ati | 0.14 | 0.13 | n/a | Yes | Yes | No | No |  |
| g | 1.0 | 0.02 | n/a | 0 | 0 | 0 | 0 |  |
| cg | 0.31 | 0.12 | n/a | 0 | 0 | 0 | 0 |  |
| mm | 0.07 | 0.35 | n/a | 0 | 0 | 0 | 0 |  |
| i | 0.65 | 0.22 | n/a | 0 | 2 | 1 | 1 |  |
| t | 0.36 | 0.06 | n/a | 1 | 1 | 1 | 0 |  |
| ti | 0.40 | 0.15 | n/a | n/a | 70% | 10% | 20% |  |
| ifta | 0.04 | 0.14 | n/a | n/a | 30% | 10% | 30% |  |
| ci | n/a | n/a | n/a | 1 | 2 | 1 | 2 |  |
| ct | n/a | n/a | n/a | 1 | 2 | 1 | 2 |  |
| i-ifta | 0.39 | 0.46 | n/a | n/a | 1 | 2 | 2 |  |
| t-ifta | 0.39 | 0.08 | n/a | n/a | 1 | 1 | 1 |  |
| ptc | 1.0 | 0.18 | n/a | 0 | 2 | 0 | 0 |  |
| v | 0.79 | 0.12 | n/a | 1 | 1 | 1 | 0 |  |
| cv | 0.15 | 0.37 | n/a | 0 | 1 | 1 | 1 |  |
| ah | 0.15 | 0.78 | n/a | 0 | 0 | 0 | 0 |  |

**Extended Data Table 9.** In-depth analysis of top 10 validation set cases with the highest discrepancy between local pathologist assessment and BanffNET scores across lesions (Case 2).

| Lesion | BanffNET H&E | BanffNET Silver | BanffNET PAS | Local Path | Central Path 1 | Central Path 2 | Central Path 3 | Comments |
| --- | --- | --- | --- | --- | --- | --- | --- | --- |
| tma | 0.25 | 0.95 | 0.94 | n/a | No | No | No | - Urinary tract infection<br>- MMDx (molecular analysis): ciprob 0.72, ctprob 0.41, cgprob 0.73, gprob 0.76, iprob 0.13, tprob 0.19, ahprob 0.70, ptcprob 0.60 |
| ati | 0.12 | 0.24 | 0.75 | n/a | Yes | Yes | Yes |  |
| g | 0.90 | 1.0 | 0.67 | 1 | 1 | 0 | 2 |  |
| cg | 0.75 | 0.23 | 0.49 | 3 | 1 | 3 | 2 |  |
| mm | 0.34 | 0.13 | 0.08 | 1 | 0 | 0 | 0 |  |
| i | 0.67 | 0.52 | 0.60 | 0 | 1 | 0 | 2 |  |
| t | 0.64 | 0.33 | 0.96 | 0 | 1 | 0 | 0 |  |
| ti | 0.61 | 0.63 | 0.56 | 2 | 70% | 70% | 60% |  |
| ifta | 0.57 | 1.0 | 0.78 | n/a | 70% | 70% | 50% |  |
| ci | n/a | n/a | n/a | 3 | 3 | 3 | 2 |  |
| ct | n/a | n/a | n/a | 3 | 3 | 3 | 3 |  |
| i-ifta | 0.74 | 0.92 | 0.94 | 1 | 3 | 3 | 3 |  |
| t-ifta | 0.68 | 0.59 | 0.98 | 0 | 1 | 3 | 2 |  |
| ptc | 0.16 | 0.27 | 0.32 | 0 | 0 | 0 | 2 |  |
| v | 0.12 | 0.12 | 0.12 | 0 | n/a | 0 | 1 |  |
| cv | 0.21 | 0.65 | 0.31 | 0 | n/a | n/a | n/a |  |
| ah | 1.0 | 0.33 | 0.90 | 3 | 1 | 3 | 1 |  |

**Extended Data Table 10.** In-depth analysis of top 10 validation set cases with the highest discrepancy between local pathologist assessment and BanffNET scores across lesions (Case 3).

| Lesion | BanffNET H&E | BanffNET Silver | BanffNET PAS | Local Path | Central Path 1 | Central Path 2 | Central Path 3 | Comments |
| --- | --- | --- | --- | --- | --- | --- | --- | --- |
| tma | 0.12 | 0.13 | 0.19 | Yes | No | No | No | - Urinary tract infection<br>- Almost all glomeruli fibrotic<br>- Low quality stainings |
| ati | 0.13 | 0.68 | 0.31 | No | Yes | Yes | No |  |
| g | 0.73 | 0.42 | 0.30 | 0 | 0 | 0 | 0 |  |
| cg | 0.21 | 0.12 | 0.14 | 0 | 0 | 0 | 0 |  |
| mm | 0.16 | 0.2 | 0.77 | 0 | 0 | 0 | 0 |  |
| i | 1.0 | 0.62 | 1.0 | 0 | 2 | 0 | 3 |  |
| t | 0.72 | 0.71 | 0.95 | 0 | 1 | 0 | 2 |  |
| ti | 1.0 | 0.82 | 0.95 | 0 | 90% | 90% | 90% |  |
| ifta | 0.63 | 0.81 | 0.89 | n/a | 80% | 90% | 90% |  |
| ci | n/a | n/a | n/a | n/a | 3 | 3 | 3 |  |
| ct | n/a | n/a | n/a | n/a | 3 | 3 | 3 |  |
| i-ifta | 0.56 | 0.90 | 1.0 | 0 | 3 | 3 | 3 |  |
| t-ifta | 0.94 | 0.62 | 0.75 | 0 | 1 | 3 | 3 |  |
| ptc | 0.93 | 0.18 | 0.14 | 0 | 0 | 0 | 0 |  |
| v | 0.14 | 0.12 | 0.15 | 0 | 0 | 0 | 0 |  |
| cv | 0.73 | 0.33 | 0.39 | 0 | 2 | 2 | 0 |  |
| ah | 0.84 | 0.18 | 0.53 | 0 | 2 | 2 | 0 |  |

**Extended Data Table 11.** In-depth analysis of top 10 validation set cases with the highest discrepancy between local pathologist assessment and BanffNET scores across lesions (Case 4).

| Lesion | BanffNET H&E | BanffNET Silver | BanffNET PAS | Local Path | Central Path 1 | Central Path 2 | Central Path 3 | Comments |
| --- | --- | --- | --- | --- | --- | --- | --- | --- |
| tma | 0.24 | 0.48 | 0.17 | n/a | No | No | No | - Poor quality of PAS staining<br>- MMDx (molecular analysis): ciprob 0.88, ctprob 0.82, cgprob 0.89, gprob 0.89, iprob 0.19, tprob 0.15, ahprob 0.87. ptcprob 0.75 |
| ati | 0.12 | 0.13 | 0.14 | n/a | No | No | No |  |
| g | 1.0 | 0.68 | 0.61 | 2 | 1 | 0 | 1 |  |
| cg | 0.36 | 0.15 | 0.72 | 0 | 1 | 1 | 2 |  |
| mm | 0.69 | 0.47 | 0.16 | 0 | 0 | 0 | 0 |  |
| i | 0.40 | 0.77 | 0.25 | 1 | 0 | 0 | 3 |  |
| t | 0.17 | 0.23 | 0.39 | 0 | 1 | 0 | 1 |  |
| ti | 0.47 | 0.67 | 0.32 | 0 | 20% | 80% | 70% |  |
| ifta | 0.24 | 0.58 | 0.66 | n/a | 50% | 80% | 40% |  |
| ci | n/a | n/a | n/a | 1 | 2 | 3 | 2 |  |
| ct | n/a | n/a | n/a | 1 | 2 | 3 | 2 |  |
| i-ifta | 0.51 | 1.0 | 0.74 | 0 | 2 | 3 | 0 |  |
| t-ifta | 0.28 | 0.37 | 0.74 | 0 | 1 | 1 | 1 |  |
| ptc | 0.30 | 1.0 | 1.0 | 3 | 3 | 3 | 2 |  |
| v | 0.12 | 0.12 | 0.12 | n/a | 0 | 0 | 0 |  |
| cv | 0.13 | 0.17 | 0.13 | n/a | 1 | 1 | 0 |  |
| ah | 0.30 | 0.13 | 0.32 | 2 | 0 | 1 | 0 |  |

**Extended Data Table 12.** In-depth analysis of top 10 validation set cases with the highest discrepancy between local pathologist assessment and BanffNET scores across lesions (Case 5).

| Lesion | BanffNET H&E | BanffNET Silver | BanffNET PAS | Local Path | Central Path 1 | Central Path 2 | Central Path 3 | Comments |
| --- | --- | --- | --- | --- | --- | --- | --- | --- |
| tma | n/a | 0.24 | 0.65 | No | Yes | No | Yes | - Recurrence of proliferative glomerulonephritis<br>- No H&E available |
| ati | n/a | 0.12 | 0.18 | Yes | No | No | No |  |
| g | n/a | 0.67 | 0.71 | 0 | 3 | 3 | 0 |  |
| cg | n/a | 0.87 | 0.38 | 3 | 3 | 3 | 3 |  |
| mm | n/a | 0.64 | 0.28 | 0 | 1 | 1 | 2 |  |
| i | n/a | 0.16 | 0.08 | 0 | 0 | 0 | 1 |  |
| t | n/a | 0.10 | 0.24 | 0 | 0 | 0 | 0 |  |
| ti | n/a | 0.15 | 0.17 | n/a | 10% | 10% | 20% |  |
| ifta | n/a | 0.27 | 0.12 | n/a | 10% | 10% | 20% |  |
| ci | n/a | n/a | n/a | 1 | 1 | 1 | 1 |  |
| ct | n/a | n/a | n/a | 1 | 1 | 1 | 1 |  |
| i-ifta | n/a | 0.87 | 0.83 | n/a | 2 | 3 | 2 |  |
| t-ifta | n/a | 0.22 | 0.5 | n/a | 0 | 1 | 0 |  |
| ptc | n/a | 0.44 | 0.57 | 0 | 0 | 0 | 0 |  |
| v | n/a | 0.12 | 0.14 | 0 | 0 | 0 | 0 |  |
| cv | n/a | 0.25 | 0.88 | 0 | 1 | 1 | 2 |  |
| ah | n/a | 0.46 | 0.58 | 0 | 1 | 2 | 2 |  |

**Extended Data Table 13.** In-depth analysis of top 10 validation set cases with the highest discrepancy between local pathologist assessment and BanffNET scores across lesions (Case 6).

| Lesion | BanffNET H&E | BanffNET Silver | BanffNET PAS | Local Path | Central Path 1 | Central Path 2 | Central Path 3 | Comments |
| --- | --- | --- | --- | --- | --- | --- | --- | --- |
| tma | n/a | 0.13 | 0.99 | No | No | No | No | - H&E staining is labeled as PAS<br>- No PAS available<br>- BanffNET H&E model applied to the wrongly labeled PAS staining: tma 0.15, ati 1.0, g 0.13, cg 0.24, mm 0.00, i 0.05, t 0.49, ti 0.01, ifta 0.00, i-ifta 0.01, t-ifta 0.04, ptc 0.27, v 0.94, cv 1.0, ah 0.98. |
| ati | n/a | 0.12 | 1.0 | Yes | No | No | Yes |  |
| g | n/a | 0.03 | 0.04 | 0 | 0 | 0 | 0 |  |
| cg | n/a | 0.12 | 0.97 | 0 | 0 | 0 | 0 |  |
| mm | n/a | 0.36 | 0.14 | 0 | 0 | 0 | 0 |  |
| i | n/a | 0.09 | 0.02 | 0 | 0 | 0 | 0 |  |
| t | n/a | 0.26 | 0.08 | 1 | 1 | 0 | 0 |  |
| ti | n/a | 0.03 | 0.02 | n/a | 0% | 10% | 10% |  |
| ifta | n/a | 0.02 | 0.00 | n/a | 0% | 0% | 10% |  |
| ci | n/a | n/a | n/a | 0 | 0 | 0 | 1 |  |
| ct | n/a | n/a | n/a | 1 | 1 | 0 | 1 |  |
| i-ifta | n/a | 0.28 | 0.00 | n/a | 0 | 0 | 0 |  |
| t-ifta | n/a | 0.36 | 0.00 | n/a | 0 | 0 | 0 |  |
| ptc | n/a | 0.14 | 1.0 | 3 | 0 | 0 | 0 |  |
| v | n/a | 0.12 | 0.18 | 0 | 2 | 2 | 0 |  |
| cv | n/a | 0.16 | 1.0 | 0 | 0 | 0 | 0 |  |
| ah | n/a | 0.67 | 1.0 | 1 | 0 | 0 | 1 |  |

**Extended Data Table 14.**
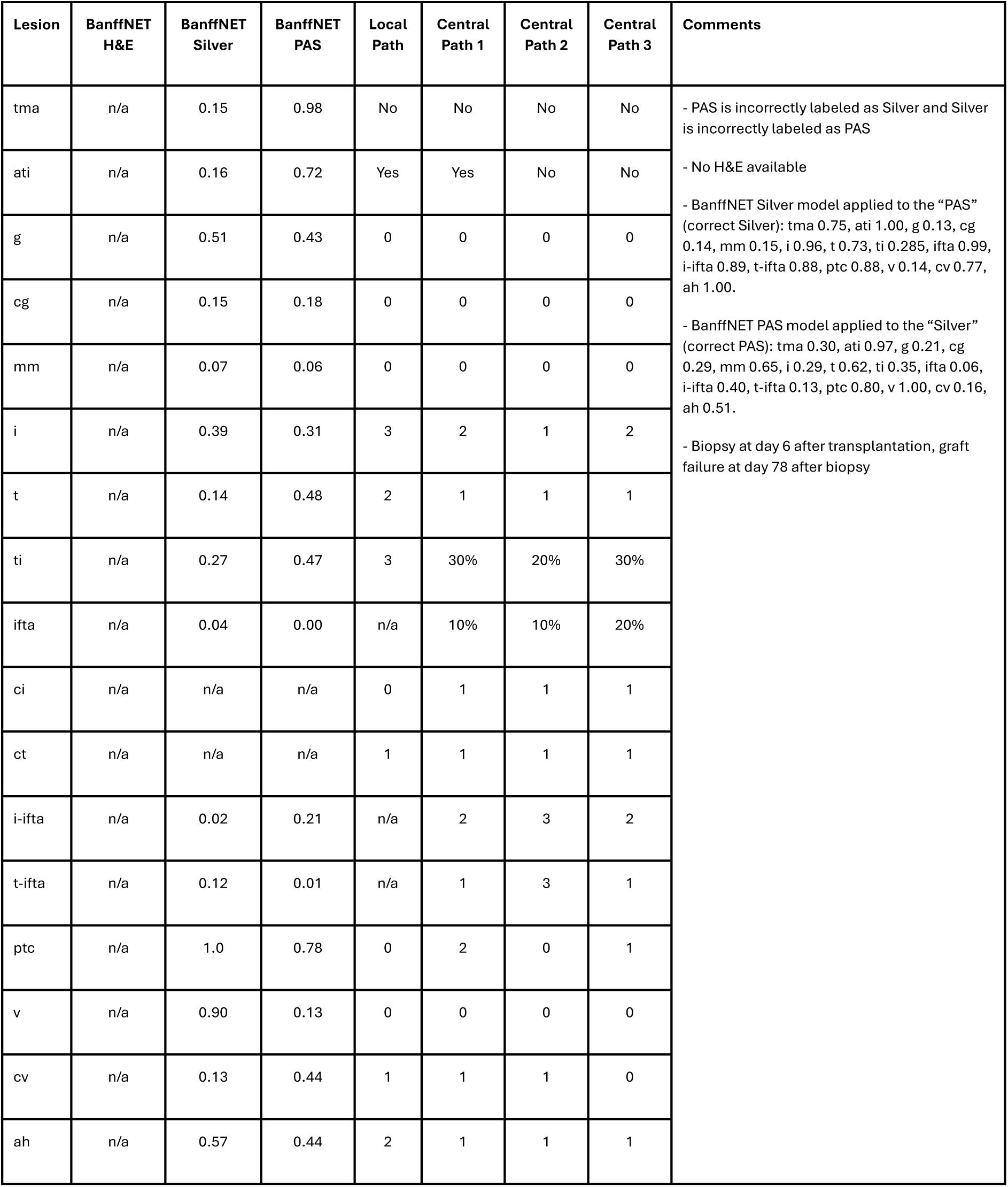
In-depth analysis of top 10 validation set cases with the highest discrepancy between local pathologist assessment and BanffNET scores across lesions (Case 7).

| Lesion | BanffNET H&E | BanffNET Silver | BanffNET PAS | Local Path | Central Path 1 | Central Path 2 | Central Path 3 | Comments |
| --- | --- | --- | --- | --- | --- | --- | --- | --- |
| tma | n/a | 0.15 | 0.98 | No | No | No | No | - PAS is incorrectly labeled as Silver and Silver is incorrectly labeled as PAS<br>- No H&E available<br>- BanffNET Silver model applied to the "PAS" (correct Silver): tma 0.75, ati 1.00, g 0.13, cg 0.14, mm 0.15, i 0.96, t 0.73, ti 0.285, ifta 0.99, i-ifta 0.89, t-ifta 0.88, ptc 0.88, v 0.14, cv 0.77, ah 1.00.<br>- BanffNET PAS model applied to the "Silver" (correct PAS): tma 0.30, ati 0.97, g 0.21, cg 0.29, mm 0.65, i 0.29, t 0.62, ti 0.35, ifta 0.06, i-ifta 0.40, t-ifta 0.13, ptc 0.80, v 1.00, cv 0.16, ah 0.51.<br>- Biopsy at day 6 after transplantation, graft failure at day 78 after biopsy |
| ati | n/a | 0.16 | 0.72 | Yes | Yes | No | No |  |
| g | n/a | 0.51 | 0.43 | 0 | 0 | 0 | 0 |  |
| cg | n/a | 0.15 | 0.18 | 0 | 0 | 0 | 0 |  |
| mm | n/a | 0.07 | 0.06 | 0 | 0 | 0 | 0 |  |
| i | n/a | 0.39 | 0.31 | 3 | 2 | 1 | 2 |  |
| t | n/a | 0.14 | 0.48 | 2 | 1 | 1 | 1 |  |
| ti | n/a | 0.27 | 0.47 | 3 | 30% | 20% | 30% |  |
| ifta | n/a | 0.04 | 0.00 | n/a | 10% | 10% | 20% |  |
| ci | n/a | n/a | n/a | 0 | 1 | 1 | 1 |  |
| ct | n/a | n/a | n/a | 1 | 1 | 1 | 1 |  |
| i-ifta | n/a | 0.02 | 0.21 | n/a | 2 | 3 | 2 |  |
| t-ifta | n/a | 0.12 | 0.01 | n/a | 1 | 3 | 1 |  |
| ptc | n/a | 1.0 | 0.78 | 0 | 2 | 0 | 1 |  |
| v | n/a | 0.90 | 0.13 | 0 | 0 | 0 | 0 |  |
| cv | n/a | 0.13 | 0.44 | 1 | 1 | 1 | 0 |  |
| ah | n/a | 0.57 | 0.44 | 2 | 1 | 1 | 1 |  |

**Extended Data Table 15.**
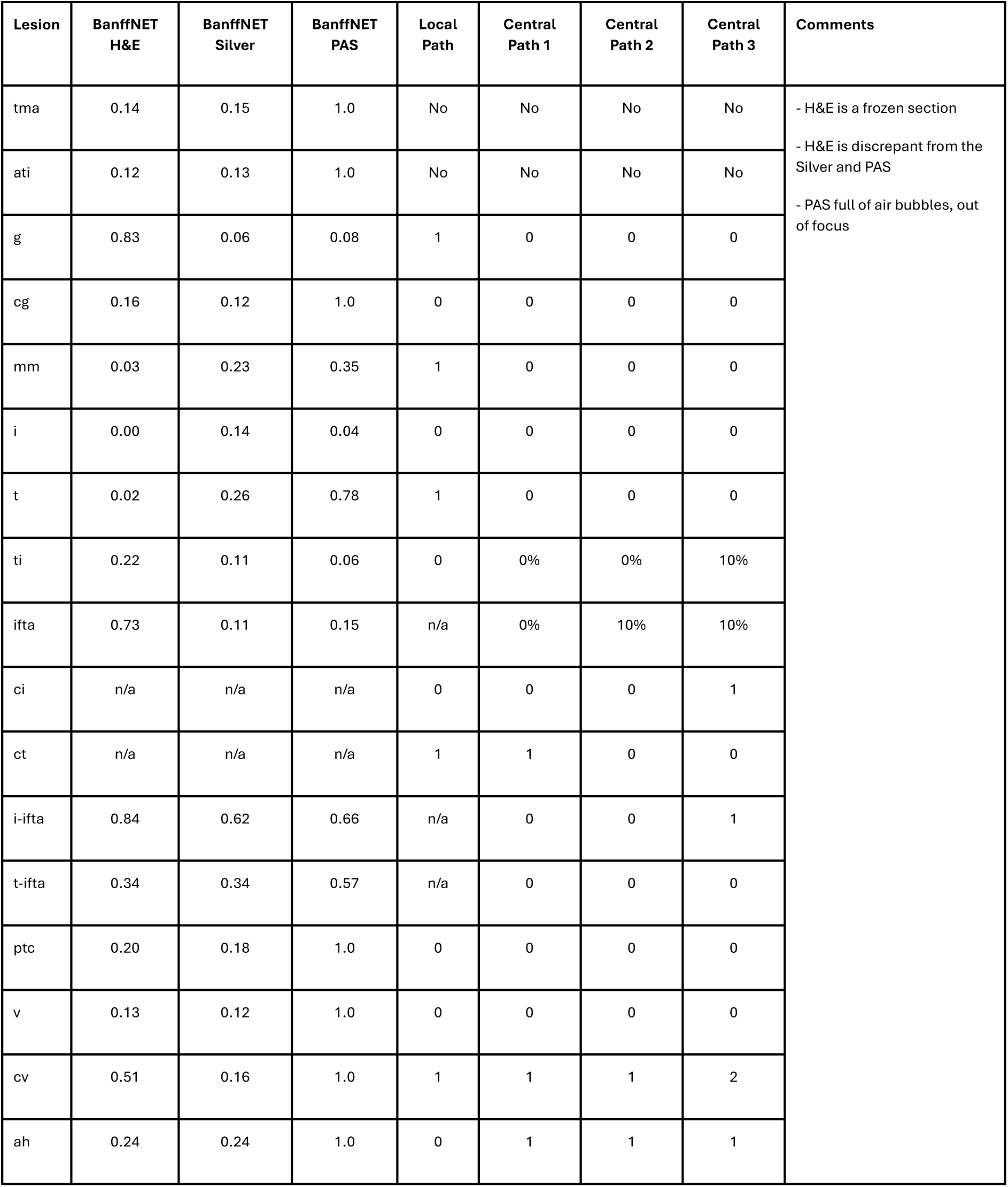
In-depth analysis of top 10 validation set cases with the highest discrepancy between local pathologist assessment and BanffNET scores across lesions (Case 8).

**Extended Data Table 16.** In-depth analysis of top 10 validation set cases with the highest discrepancy between local pathologist assessment and BanffNET scores across lesions (Case 9).

| Lesion | BanffNET H&E | BanffNET Silver | BanffNET PAS | Local Path | Central Path 1 | Central Path 2 | Central Path 3 | Comments |
| --- | --- | --- | --- | --- | --- | --- | --- | --- |
| tma | 0.37 | 0.84 | 0.86 | No | No | No | No | - Recurrence of proliferative glomerulonephritis<br>- Extensive interstitial fibrosis and tubular atrophy |
| ati | 0.26 | 0.38 | 0.58 | No | No | No | No |  |
| g | 1.0 | 0.75 | 1.0 | 0 | 2 | 3 | 3 |  |
| cg | 1.0 | 1.0 | 1.0 | 3 | 3 | 3 | 3 |  |
| mm | 0.67 | 0.38 | 0.76 | 0 | 2 | 3 | 1 |  |
| i | 0.40 | 0.79 | 0.48 | 3 | 1 | 0 | 3 |  |
| t | 0.20 | 0.50 | 0.08 | 1 | 0 | 0 | 0 |  |
| ti | 0.83 | 0.76 | 0.53 | 3 | 40% | 100% | 90% |  |
| ifta | 0.68 | 0.74 | 0.79 | n/a | 80% | 100% | 90% |  |
| ci | n/a | n/a | n/a | 3 | 3 | 3 | 3 |  |
| ct | n/a | n/a | n/a | 1 | 3 | 3 | 3 |  |
| i-ifta | 0.73 | 0.64 | 0.38 | n/a | 1 | 3 | 2 |  |
| t-ifta | 0.21 | 0.65 | 0.24 | n/a | 1 | 1 | 0 |  |
| ptc | 1.0 | 1.0 | 0.99 | 0 | 0 | 3 | 0 |  |
| v | 0.15 | 0.25 | 0.27 | 0 | 0 | 0 | 0 |  |
| cv | 0.28 | 0.19 | 0.15 | 0 | 0 | 1 | 0 |  |
| ah | 0.65 | 0.39 | 0.98 | 0 | 1 | 1 | 0 |  |

**Extended Data Table 17.** In-depth analysis of top 10 validation set cases with the highest discrepancy between local pathologist assessment and BanffNET scores across lesions (Case 10).

| Lesion | BanffNET H&E | BanffNET Silver | BanffNET PAS | Local Path | Central Path 1 | Central Path 2 | Central Path 3 | Comments |
| --- | --- | --- | --- | --- | --- | --- | --- | --- |
| tma | 0.12 | 0.24 | 0.39 | No | No | No | No | - Recurrence of membranous glomerulopathy |
| ati | 0.25 | 0.21 | 0.84 | Yes | No | No | Yes |  |
| g | 0.34 | 0.66 | 0.62 | 0 | 1 | 3 | 1 |  |
| cg | 0.97 | 0.92 | 0.61 | 3 | 1 | 2 | 1 |  |
| mm | 0.42 | 0.33 | 0.40 | 3 | 0 | 3 | 0 |  |
| i | 0.29 | 0.20 | 0.66 | 3 | 0 | 0 | 3 |  |
| t | 0.70 | 0.46 | 0.58 | 1 | 1 | 0 | 0 |  |
| ti | 0.82 | 0.60 | 0.68 | 3 | 50% | 70% | 70% |  |
| ifta | 0.46 | 0.52 | 0.75 | n/a | 60% | 70% | 60% |  |
| ci | n/a | n/a | n/a | 1 | 3 | 3 | 2 |  |
| ct | n/a | n/a | n/a | 1 | 3 | 3 | 3 |  |
| i-ifta | 0.95 | 0.90 | 0.84 | 3 | 2 | 3 | 3 |  |
| t-ifta | 0.91 | 0.46 | 0.51 | 1 | 1 | 3 | 3 |  |
| ptc | 0.5 | 0.66 | 0.65 | 0 | 0 | 0 | 1 |  |
| v | 0.12 | 0.14 | 0.14 | 0 | 0 | 0 | 0 |  |
| cv | 0.94 | 0.67 | 0.73 | 1 | 1 | 1 | 1 |  |
| ah | 0.99 | 1.0 | 1.0 | 2 | 2 | 3 | 3 |  |

**Extended Data Table 18.**
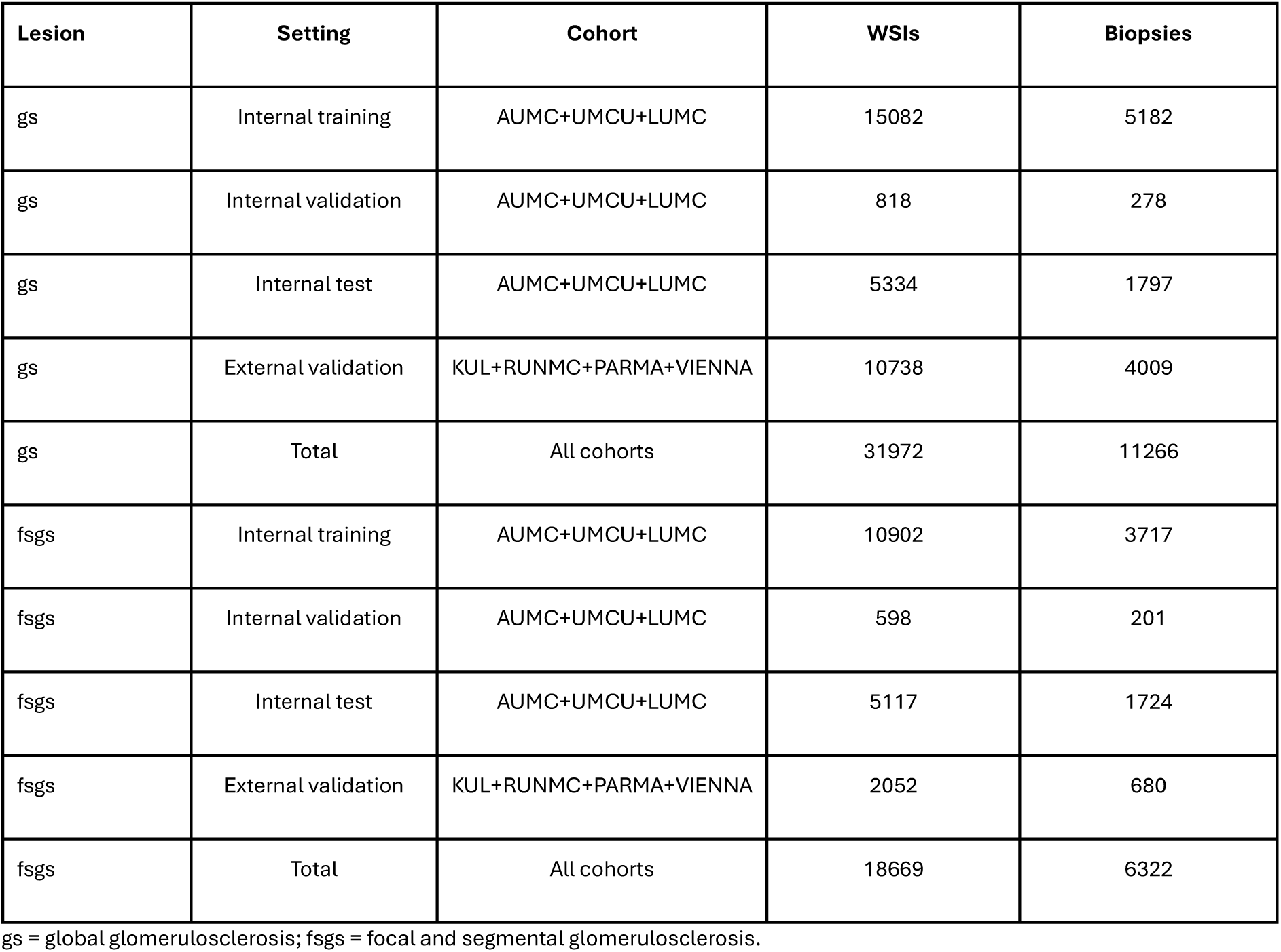
Breakdown of whole slide images for the training, validation and testing of global glomerulosclerosis and focal and segmental glomerulosclerosis.

| Lesion | Setting | Cohort | WSIs | Biopsies |
| --- | --- | --- | --- | --- |
| gs | Internal training | AUMC+UMCU+LUMC | 15082 | 5182 |
| gs | Internal validation | AUMC+UMCU+LUMC | 818 | 278 |
| gs | Internal test | AUMC+UMCU+LUMC | 5334 | 1797 |
| gs | External validation | KUL+RUNMC+PARMA+VIENNA | 10738 | 4009 |
| gs | Total | All cohorts | 31972 | 11266 |
| fsgs | Internal training | AUMC+UMCU+LUMC | 10902 | 3717 |
| fsgs | Internal validation | AUMC+UMCU+LUMC | 598 | 201 |
| fsgs | Internal test | AUMC+UMCU+LUMC | 5117 | 1724 |
| fsgs | External validation | KUL+RUNMC+PARMA+VIENNA | 2052 | 680 |
| fsgs | Total | All cohorts | 18669 | 6322 |

gs = global glomerulosclerosis; fsgs = focal and segmental glomerulosclerosis.

