## Supplementary material for "BanffNET, a Deep Learning System for Comprehensive Histological Lesion Quantification in Kidney Transplant Biopsies": CLAIM checklist

### Checklist for Artificial Intelligence in Medical Imaging (CLAIM): 2024 Update

| Section / Topic | No. | Item | Page / Line | No | NA |
|---|---|---|---|---|---|
| **TITLE / ABSTRACT** | | | | | |
| | **1** | Identification as a study of AI methodology, specifying the category of technology used (e.g., deep learning) | **1,6** | | |
| **ABSTRACT** | | | | | |
| | **2** | Summary of study design, methods, results, and conclusions | **6** | | |
| **INTRODUCTION** | | | | | |
| | **3** | Scientific and/or clinical background, including the intended use and role of the AI approach | **7** | | |
| | **4** | Study aims, objectives, and hypotheses | **7** | | |
| **METHODS** | | | | | |
| ***Study Design*** | **5** | Prospective or retrospective study | **23-25, 29-30** | | |
| | **6** | Study goal | **7, 27-28** | | |
| ***Data*** | **7** | Data sources | **7-8, 22-25** | | |
| | **8** | Inclusion and exclusion criteria | **23-25, 26** | | |
| | **9** | Data pre-processing | **26-27** | | |
| | **10** | Selection of data subsets | **7-8, 23-26, 29** | | |
| | **11** | De-identification methods | **24-25** | | |
| | **12** | How missing data were handled | **28** | | |
| | **13** | Image acquisition protocol | **23-25, 26** | | |
| ***Reference Standard*** | **14** | Definition of method(s) used to obtain reference standard | **8, 25-26, 27** | | |
| | **15** | Rationale for choosing the reference standard | **7-8, 20-21** | | |
| | **16** | Source of reference standard annotations | **7-8, 23-26** | | |
| | **17** | Annotation of test set | **24-26** | | |
| | **18** | Measures of inter- and intra-rater variability of features described by the annotators | **7, 12-14, 25-26** | | |
| ***Data Partitions*** | **19** | How data were assigned to partitions | **7-8, 29** | | |
| | **20** | Level at which partitions are disjoint | **7-8, 23-25** | | |
| ***Testing Data*** | **21** | Intended sample size | **7-8, 47** | | |

| Section / Topic | No. | Item | Page / Line | No | NA |
|---|---|---|---|---|---|
| ***Model*** | **22** | Detailed description of model | **8-9, 26-28, 50** | | |
| | **23** | Software libraries, frameworks, and packages | **29-30** | | |
| | **24** | Initialization of model parameters | **26, 28** | | |
| ***Training*** | **25** | Details of training approach | **27-28** | | |
| | **26** | Method of selecting the final model | **28-29** | | |
| | **27** | Ensembling techniques | **8-9, 27** | | |
| ***Evaluation*** | **28** | Metrics of model performance | **27-29** | | |
| | **29** | Statistical measures of significance and uncertainty | **28-29** | | |
| | **30** | Robustness or sensitivity analysis | **11, 53-63** | | |
| | **31** | Methods for explainability or interpretability | **9, 28, 35** | | |
| | **32** | Evaluation on internal data | **10-11, 16, 36, 42** | | |
| | **33** | Testing on external data | **10-20, 37, 42-46** | | |
| | **34** | Clinical trial registration | **24** | | |
| **RESULTS** | | | | | |
| ***Data*** | **35** | Numbers of patients or examinations included and excluded | **7-8, 23-26, 47, 64** | | |
| | **36** | Demographic and clinical characteristics of cases in each partition | **47-48** | | |
| ***Model performance*** | **37** | Performance metrics and measures of statistical uncertainty | **10-20, 52** | | |
| | **38** | Estimates of diagnostic performance and their precision | **10, 13, 15, 17, 19** | | |
| | **39** | Failure analysis of incorrect results | **11, 38-41, 53-63** | | |
| **DISCUSSION** | | | | | |
| | **40** | Study limitations | **22** | | |
| | **41** | Implications for practice, including intended use and/or clinical role | **21-22** | | |
| **OTHER INFORMATION** | | | | | |
| | **42** | Provide a reference to the full study protocol or to additional technical details | **24, 26-29, 65-67** | | |

| | | | |
|---|---|---|---|
| | **43** | Statement about the availability of software, trained model, and/or data | **30** |
| | **44** | Sources of funding and other support; role of funders | **30-31** |

* Indicate page and/or line number for each checklist item that is present.  NA = not applicable.
